# A multi-level gene-diet interaction analysis of fish oil supplementation and 14 circulating polyunsaturated fatty acids-related traits identifies the *FADS* and *GRP12* loci

**DOI:** 10.1101/2024.12.12.24318956

**Authors:** Susan Adanna Ihejirika, Alexandra Huong Chiang, Aryaman Singh, Eunice Stephen, Han Chen, Kaixiong Ye

## Abstract

Fish oil supplements (FOS) are known to alter circulating levels of polyunsaturated fatty acids (PUFAs) among individuals but in a heterogeneous manner. These varied responses may result from unidentified gene-FOS interactions. To identify genetic factors that interact with FOS to alter the circulating levels of PUFAs, we performed a multi-level genome-wide interaction study (GWIS) of FOS on 14 plasma measurements in 200,060 unrelated European-ancestry individuals from the UK Biobank. From our single-variant tests, we identified genome-wide significant interacting SNPs (*P* < 5 × 10^−8^) in the *FADS1-FADS2* gene cluster for total omega-3, omega-3%, docosapentaenoic acid (DHA), DHA% and the omega-6 to omega-3 ratio. Among the interaction signals for omega-3%, the lead SNP, rs35473591 (C>CT, CT allele frequency = 0.34), had a lower association effect size in the FOS-taking group (β = 0.35 for allele C) than that in the group without FOS (β = 0.42). Likewise, the effect sizes of associations between FOS and omega-3% varied across the three genotype groups (β = 0.45, 0.50, and 0.59, respectively, in C/C, C/CT, and CT/CT). Our gene-level aggregate and transcriptome-wide interaction analyses identified significant signals at two loci, around *FADS1-FADS2* and *GRP12*. The contribution of genome-wide gene-FOS interactions to phenotypic variance was statistically significant in omega-3-related traits. This systemic gene-FOS GWIS contributes to our understanding of the genetic architecture of circulating PUFAs underlying FOS response and informs personalized dietary recommendations.

## Introduction

Genetic^1–3^ and environmental factors, such as diet^4–6^, influence the circulating levels of polyunsaturated fatty acids (PUFAs). PUFAs are fatty acids with 18 to 24 carbons and multiple double bonds. They fall into two main families, omega-3 and omega-6, depending on which carbon the first double bond falls on from the methyl end of the carbon chain. PUFAs play crucial roles in various physiological processes, including immune function, cardiovascular health, brain development, and cognitive performance.^7–9^ Alpha-linolenic acid (ALA) and linoleic acid (LA) are essential omega-3 and omega-6 fatty acids, respectively, that can only be obtained from dietary sources and act as precursors for synthesizing long-chain PUFAs.^7^ Long-chain PUFAs can be obtained from diet or produced endogenously in the presence of the necessary precursors.^7^ Fish oil supplements (FOS) are a rich, readily available source of long-chain omega-3 PUFAs, particularly eicosapentaenoic acid (EPA) and docosapentaenoic acid (DHA).^10^ Additionally, FOS intake alters circulating PUFA concentrations similarly to fish consumption.^11^

The interactions between genetic variants and dietary PUFAs also influence circulating PUFA concentrations.^7,8,12–19^ This means that omega-3 PUFAs obtained through FOS can alter the genetic effects on circulating PUFA levels, and that genetic variants can modify the effects of FOS on the same phenotypes. One of the relevant genetic loci, the *FADS* locus, contains the *FADS1* and *FADS2* genes, which encode the delta-5 (D5) and delta-6 (D6) desaturase enzymes, respectively. These membrane-bound enzymes introduce cis double bonds at the D5 and D6 positions in PUFAs, the rate-limiting steps in long-chain PUFA biosynthesis.^20,21^ Juan *et al.* found that higher dietary EPA and DHA consumption increases circulating EPA proportions.^12^ However, genotypes at the rs174546 *FADS1* SNP (C>T) impacted the extent of that increase. For every 1-standard deviation (SD) increase in EPA intake, individuals with the C/C genotype saw an average increase of 3.7% in the circulating EPA level, while those with the T/T genotype saw an average increase of 7.8%. These gene-diet interactions may explain some of the heterogeneity in circulating PUFA levels in response to omega-3 PUFA supplementation.^22^ Furthermore, these genotype-dependent responses may provide some insights into the inconsistent results in observational studies and randomized controlled trials (RCTs) investigating the effect of omega-3 PUFA supplementation on various outcomes,^8^ such as the risk of cardiometabolic diseases, Alzheimer’s disease, cancer, and inflammation.^23–27^

Despite advances in characterizing genetic loci interacting with dietary omega-3 PUFAs, most existing gene-dietary PUFAs interaction studies have various weaknesses and limitations. Most of them examined only candidate genes or variants^12–18^. Some were performed in specific contexts, such as disease risk and related outcomes^19,23–27^, while others only examined circulating metabolites other than PUFAs as outcomes.^19,28^ They were also limited by small sample sizes. This leaves a gap in understanding the effects of interactions between genome-wide loci and dietary omega-3 PUFAs on circulating PUFAs in a generally healthy cohort.

To this end, we conducted a large-scale gene-diet interaction analysis (N=200,060) of FOS on 14 circulating PUFAs-related measurements in the UK Biobank cohort^29,30^ to systematically identify genetic factors that interact with FOS to alter levels of plasma PUFAs. We then performed a transcriptome-wide interaction study (TWIS) to identify genes whose expression interacts with FOS. Lastly, we estimated the phenotypic variance explained by genome-wide interactions with FOS. Our study is the first large-scale GWIS of FOS on the circulating levels of PUFAs.

## Methods

### Cohort

The UK Biobank is a large, population-based prospective study of over 500,000 volunteer participants recruited between 2006 and 2010 from across England, Wales, and Scotland. The participants were between the ages of 40 and 69 years. At recruitment, participants provided sociodemographic, lifestyle, environmental, clinical, and biochemical information through touchscreen questionnaires, face-to-face interviews, and physiological measurements. Plasma samples were provided for genotyping and nuclear magnetic resonance (NMR) spectroscopy-based metabolomic measurements.^29,30^ The UK Biobank received ethical approval from the National Research Ethics Service Committee North West–Haydock (reference ID 11/NW/0382). The use of participants’ data in this study was approved under Project 48818.

### Participant Inclusion and Exclusion

In this study, we restricted our participants to those who were of genetically determined European ancestry^31^, did not have a high degree of kinship with other volunteers, were not outliers for heterozygosity or missing genotype rate, had matched self-reported and genetic sex, and had no sex chromosome aneuploidy (Figure S1). All UK Biobank participants provided written informed consent upon recruitment, authorizing the storage and use of their provided biological, medical, and genetic data for health-related research purposes. Additionally, participants who had withdrawn their consent as of February 22^nd^, 2022, were removed (N=114).

### Genotype

The genotype data used in this study was version 3 of the UK Biobank genotyping release, previously described in detail.^29^ This data included genotyping of ∼820,000 variants done with either the UK BiLEVE Axiom Array or the UKB Axiom Array. It also included initial quality control and imputation of variants using a merged reference panel of the Haplotype Reference Consortium (HRC) and UK10K.^29^ For further genotype QC, we selected only autosomal variants with imputation quality score (INFO) > 0.5, minor allele frequency (MAF) > 1%, genotype missingness per individual < 5%, genotype missingness per variant < 2%, and Hardy-Weinberg equilibrium (HWE) *p*-value < 1.0 x 10^-6^ using PLINK2 alpha-v2.3.^32^

### Phenotype

Finland-based Nightingale Health Ltd. performed NMR metabolomic measurement of 251 metabolic biomarkers for approximately 280,000 participants. The UK Biobank released this data in two phases – Phases One and Two. Phase One release consisted of plasma measurements for 120,000 randomly selected samples. Phase Two consisted of measurements for individuals in Phase One and an additional 170,000 samples. Absolute circulating levels of fatty acids, including PUFAs and monounsaturated fatty acids (MUFAs), were measured and reported in mmol/L. The proportion of each PUFA to total fatty acids was expressed as a percentage. We investigated 14 circulating PUFAs and MUFAs-related traits as our outcomes. The 14 phenotypes were the absolute circulating levels of total PUFAs, total MUFAs, omega-3 PUFAs, omega-6 PUFAs, LA, and DHA; their relative percentages in total fatty acids (i.e., PUFAs%, MUFAs%, omega-3%, omega-6%, LA%, and DHA%), and the ratios of PUFAs to MUFAs and omega-6 to omega-3. All phenotypes were rank-based inverse normal transformed.

### Dietary Exposure

The primary exposure of interest in this study is FOS. We used the UK Biobank dietary questionnaire administered during recruitment to determine the exposure status. This questionnaire was a touchscreen survey about the intake frequency of common food and drink items. A total of 497,666 participants filled these out during the initial assessment center visit between 2006 and 2010. Participants were asked about mineral and other dietary supplements, “Do you regularly take any of the following? (You can select more than one answer)”, and “fish oil (including cod liver oil)” was one of the options (Data Field 6179).

### Genome-wide Interaction Analysis

We performed single-variant interaction tests with the Gene-Environment interaction analysis in Millions of samples (GEM) tool.^33^ GEM implements a generalized linear model for unrelated individuals. GEM conducts tests for marginal effects, interaction effects (1-degree of freedom tests), and joint tests for genetic main and interaction effects (2-degree of freedom tests). In this analysis, only results from the 1-degree of freedom (1df) test were interpreted due to our interest in strong interaction effects. We used age, sex, age-by-sex, and the first ten principal components as the covariates in our model. Genome-wide significance *p-*valu*e* of 5.0 × 10^-8^ was used to define significant interactions.

To interpret the interaction signals identified by GEM, we used PLINK2 alpha-v2.3 to conduct exposure-stratified analysis to quantify SNP effects in each exposure group. Lastly, to quantify the effects of FOS across the genotype groups, we fitted linear models to each group and adjusted for the same covariates as in our single-variant interaction tests.

### Gene-level Interaction Analysis

FUMA^34^ was used to implement the MAGMA model^35^, which maps variants to genes based on physical locations, aggregates *p*-values from the individual variant-FOS interaction tests across genic regions, and tests gene-level associations with each of the phenotypes of interest. Variants were mapped to 18,767 genes based on defined transcription start and stop sites, and a *p*-value of 2.66 × 10^-6^ (0.05/18767) was used to define statistical significance. Linkage disequilibrium (LD) was estimated using the UK Biobank release 2b 10k white British reference panel.

### Transcriptome-wide Interaction Analysis

We used the MetaXcan framework for our TWIS to integrate eQTL information with single-variant GWIS summary statistics. MetaXcan tools include S-PrediXcan^36^ and S-MultiXcan.^37^ S-PrediXcan predicts and imputes genetically determined gene expression levels and tests their associations with the phenotype of interest. We used the MASHR-based models to predict gene expression. These models are based on fine-mapped variants for each tissue. They take in gene expression weights, the variance and covariances of SNPs, and the beta coefficient of each SNP from summary statistics. S-MultiXcan aggregates information across all tissues. After multiple testing correction for the number of genes that had a prediction model in at least one tissue, significance was defined as *p* < 2.9 × 10^-6^ (0.05/17,500).

### Estimation of Phenotypic Variance Explained by Gene-FOS Interactions

We performed GCTA-GREML^38^ analysis to estimate the proportion of phenotypic variance explained by genome-wide gene-FOS interactions for each of the 14 phenotypes separately. We estimated the genetic relationship matrix (GRM) between pairs of individuals using LD-pruned SNPs. SNPs were pruned using a window size of 1000 variants, a step size of 100 variants, and an r^2^ of 0.9.

To estimate the variance explained by the top variant, we assumed gene-environment independence and applied the formula

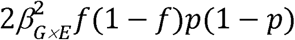

where β is the gene-FOS interaction effect size, *f* is the MAF, and *p* is the exposure prevalence in the population.

## Results

### Sample characteristics

NMR metabolite measurements for UK Biobank participants were released in two phases – Phase One (*N* = 120,000) and Phase Two (*N* = 170,000 additional samples). Phase Two release also included updated and corrected Phase One measurements (Methods). We refer to the additional 170,000 samples as Phase Two in this study. We performed exposure quality control on the combined dataset of 290,000 participants for our gene-FOS interaction analysis (Figure S1). Approximately 31.8% (*N* = 63,711) of 200,060 included participants indicated regular FOS use at recruitment (Table S1). The percentage of females and mean age in our dataset were 53.5% and 56.8 ± 8.0, respectively.

### Variant-level analysis identified interacting SNPs in the *FADS* locus

We performed single-variant 1-df interaction tests to examine the interactions between FOS and genetic variants in the Phase One and Phase Two datasets, separately. We identified one and two unique loci at *p* < 5.0 × 10^-8^ in the Phase One and Two datasets, respectively (Tables S2 and S3, Figures S2, S3, and S4). The correlations between the *p-*values from both analyses are shown in Figure S3, with reproducible association signals observed for total omega-3, omega-3%, the omega-6/omega-3 ratio, DHA, and DHA%. These reproducible signals, genome-wide significant in Phase Two and nominally significant in Phase One, locate in the *FADS1-FADS2* gene cluster, hereafter referred to as the *FADS* locus.

Next, we performed interaction tests in the combined dataset of Phase One and Two (Figure S5, Table S4). We identified genome-wide significant interaction effects for variants in the *FADS* locus, for five traits, including total omega-3, omega-3%, DHA, DHA%, and the omega-6/omega-3 ratio (Figures 1A and S5, Table 1). To report our significant results, we focused on omega-3% as we observed similar trends in the other four traits. Omega-3% encompasses all omega-3 PUFAs and reflects omega-3 concentration in relation to total fatty acids. The most significant interaction was at the lead SNP rs35473591 (chr11:61586328; C>CT, MAF = 0.34) (Figure 1B). To quantify the genetic effect of rs35473591 in each exposure subgroup, we performed a stratified analysis (Figure 1C). The C allele had an overall positive effect on omega-3%, but its effect size was lower in the FOS-taking group (β = 0.35) than in the group that does not use FOS (β = 0.42). Furthermore, we fitted linear models to quantify the effect of FOS in each genotype subgroup. FOS significantly altered omega-3% levels. The degree of alteration varied across the three genotype groups (Figure 1D, β = 0.45, 0.50, 0.59 SD units, respectively, in C/C, C/CT, and CT/CT). Together, these results highlight a significant effect of interaction between rs35473591 and FOS on omega-3%, demonstrating that the effect of FOS on circulating omega-3% is modulated by rs35473591 genotype. Similar interaction effects were observed at the *FADS* locus for total omega-3, DHA, DHA%, and the omega-6/omega-3 ratio.

**Figure 1.**
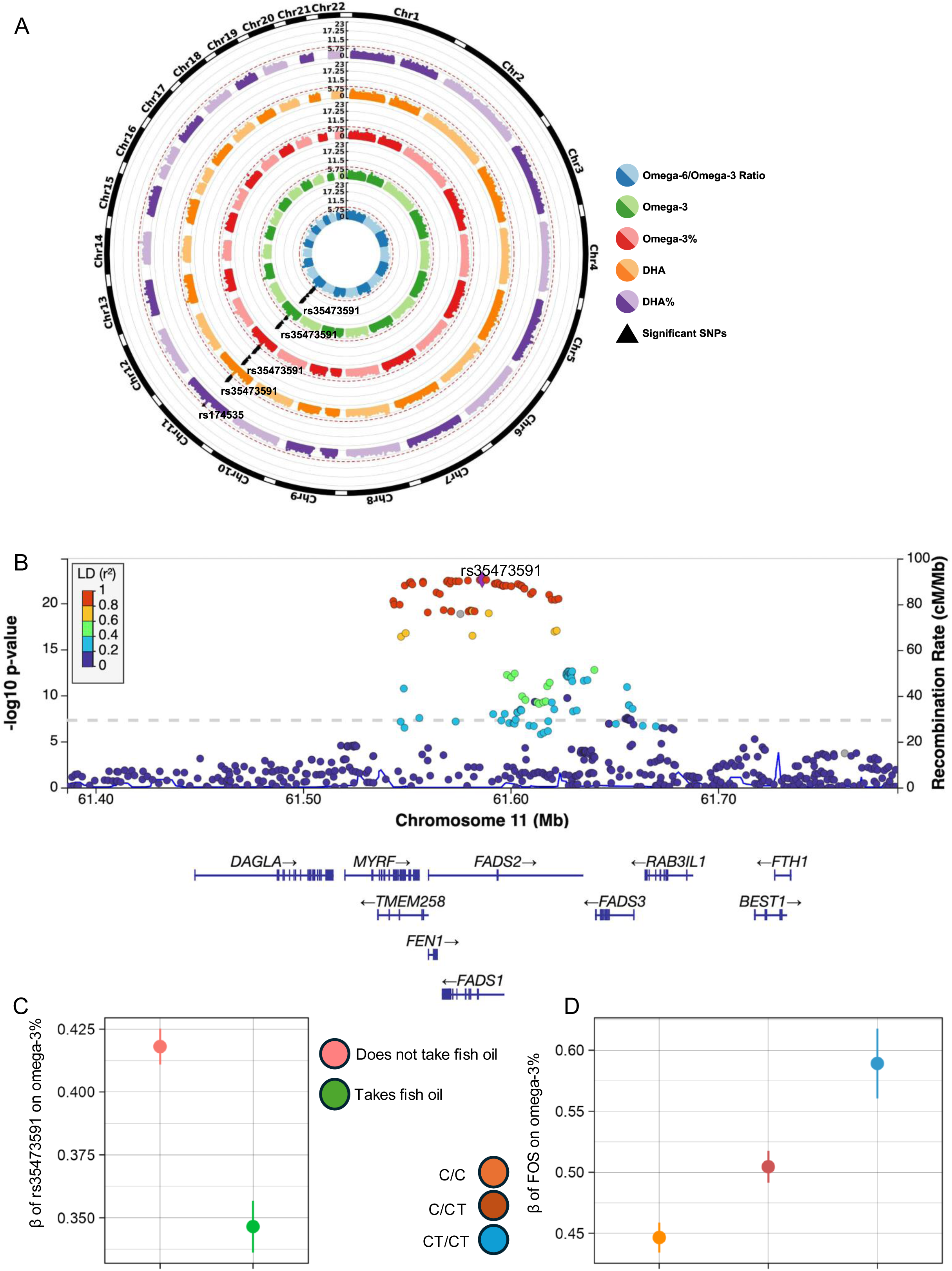
*FADS* variants with significant interaction signals (*p* < 5.0 × 10^−8^) in five PUFA traits. A) Circular Manhattan plots of *p*-values for gene-FOS interactions on omega-3, omega-3%, DHA, DHA%, and the ratio of omega-6 to omega-3. B) LocusZoom plot of the *FADS* locus for omega-3%. C) Marginal genetic effect of rs35473591 on omega-3% in stratified exposure subgroups. D) Marginal exposure effects of FOS on omega-3% in stratified genotype subgroups. All error bars represent 95% confidence intervals.

**Table 1.** Lead SNPs in traits with genome-wide significant interaction signals. All significant variants are in the *FADS1-FADS2* gene cluster.

| rsID | Chr | Pos | Nearest gene | EA<br>(frequency) | NEA | PUFA trait | Interaction<br>*p*-value | Interaction<br>Beta | Interaction<br>S.E. |
| --- | --- | --- | --- | --- | --- | --- | --- | --- | --- |
| rs35473591 | 11 | 61586328 | *FADS1*-<br>*FADS2* | C (0.66) | CT | Omega-3% | $2.50 \times 10^{-23}$ | -0.0722 | 0.00726 |
| | | | | | | Omega-6 to<br>Omega-3 ratio | $1.09 \times 10^{-20}$ | 0.0672 | 0.00720 |
| | | | | | | Omega-3 | $2.74 \times 10^{-17}$ | -0.0595 | 0.00704 |
| | | | | | | DHA | $1.56 \times 10^{-12}$ | -0.0492 | 0.00696 |
| rs174535 | 11 | 61551356 | *MYRF* | T (0.65) | C | DHA% | $4.11 \times 10^{-8}$ | -0.0390 | 0.00718 |

EA, effect allele; NEA, non-effect allele; S.E., standard error

### Gene-level interactions with FOS

To extend our single-variant tests, we first used MAGMA to aggregate interaction *p*-values across genic regions and performed gene-based interaction tests. The *FADS* cluster interaction was significant in the same five traits as in the single-variant tests (Figure 2). Around this *FADS* locus, in addition to the *FADS1* and *FADS2* genes, the *FADS3, TMEM258, MYRF, FEN1,* and *RAB3IL1* genes were also significant across the same five traits (Figure 2A), but these signals may be driven by the same variants due to the high level of LD in this genomic region. Additionally, a novel locus around the *GPR12* gene was significant in total PUFAs% and the ratio of PUFAs to MUFAs (Figure 2B). The top SNP in PUFAs%, rs1752653 (chr 13:27322777; T>C, MAF = 0.21), reached suggestive genome-wide significance at *p <* 5.0 × 10^-6^ (Figure 3A). *GPR12-*rs1752653 had opposite directions of genetic effects on PUFAs% in the FOS and non-FOS-taking groups, with β = 0.03 and −0.01, respectively (Figure 3B). Across the three genotype groups, the effect of FOS on PUFAs% is β = 0.24, 0.21, and 0.16 SD units, respectively, in T/T, T/C, and C/C (Figure 3C).

**Figure 2.**
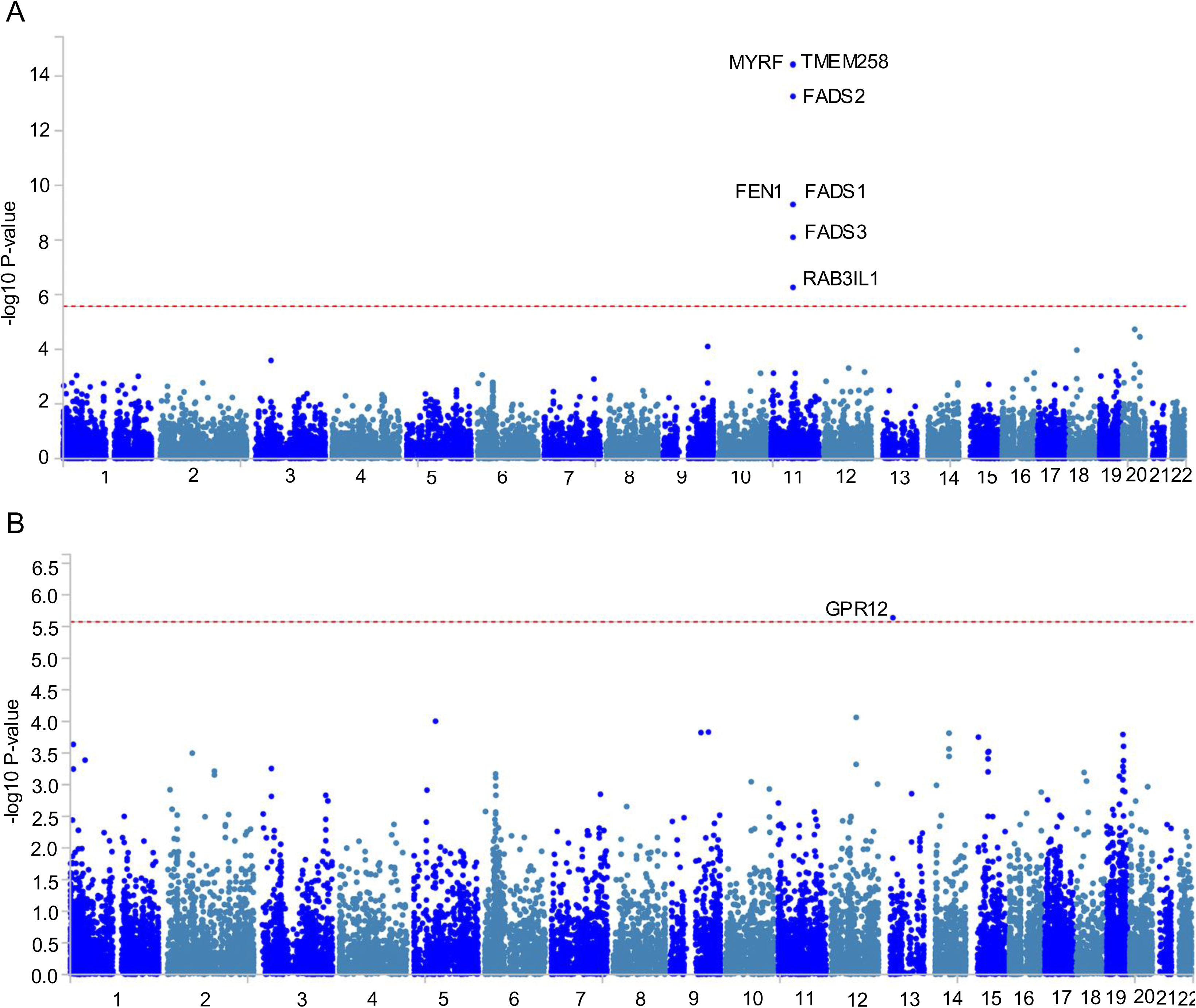
Significant gene-level interactions for Omega-3% and PUFAs%. Gene-level Manhattan plots from MAGMA analysis for genes that reached the significance threshold of *p <* 2.66 × 10^-6^ in A) Omega-3% and B) PUFAs%, C) Marginal genetic effect of rs1752653 on PUFAs% in stratified exposure subgroups. Error bars show 95% confidence intervals. D) Marginal exposure effects of FOS on PUFAs% in stratified genotype subgroups. Error bars represent 95% confidence intervals.

**Figure 3.**
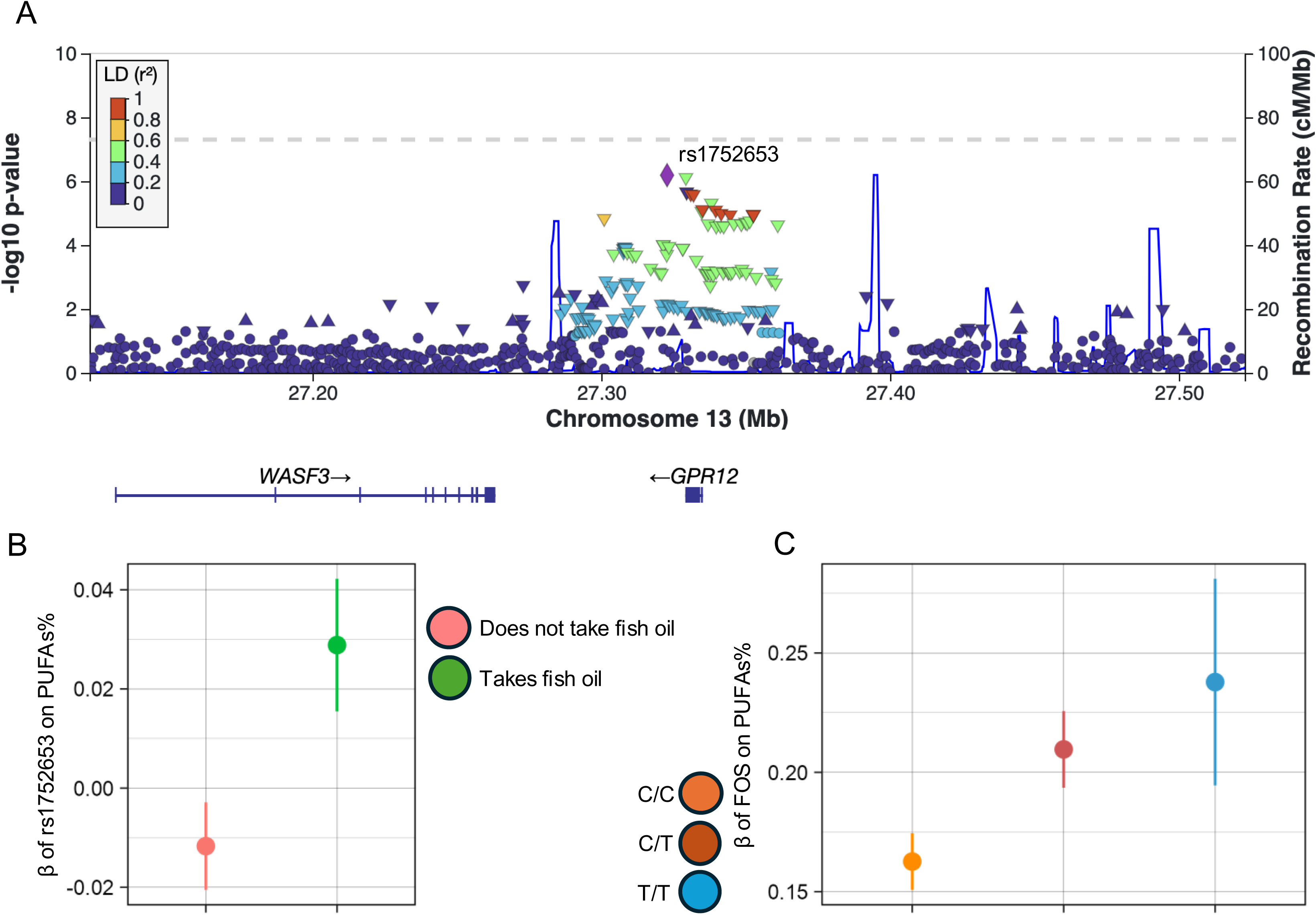
GPR12 variant with suggestive genome-wide significance (p < 5.0 × 10-6) in PUFAs%. A) LocusZoom plot of the GPR12 locus for PUFAs%. B) Marginal genetic effect of rs1752653 on PUFAs% in stratified exposure subgroups. C) Marginal exposure effects of FOS on PUFAs% in stratified genotype subgroups. All error bars represent 95% confidence intervals.

Next, we integrated eQTL information with our GWIS summary statistics using S-PrediXcan and S-MultiXcan. We first performed S-PrediXcan analyses to identify genes whose genetically predicted expression levels interact significantly with FOS intake status in each phenotype. S-PrediXcan conducted single-tissue analysis in 49 tissues available in the GTEx consortium. We used S-MultiXcan to aggregate results across multiple tissues and to increase power relative to the individual tissue analyses. After multiple testing correction for the number of genes that had a prediction model in at least one tissue (0.05/17,500; *p* < 2.9 × 10^-6^), we identified 24 significant interactions involving seven genes across five unique traits (Table S5). The *FADS1, FADS2, FADS3, TMEM258, FEN1, BEST1,* and *FTH1* genes were all significant across the same five traits as MAGMA gene-based analysis and our single-variant tests. The most significant interaction was for the *FADS1* gene (*p* = 8.8 x 10^-22^) in omega-3%. The *FADS1* gene was also the only gene with interaction effects in all five phenotypes with significant interactions. In summary, two types of gene-based analyses consistently identified genes around the *FADS* locus to exhibit significant gene-FOS interaction signals for five PUFA traits. Gene-level analysis with MAGMA further identified the *GPR12* locus for total PUFAs% and the ratio of PUFAs to MUFAs.

### Contribution of gene-FOS interactions to the phenotypic variance of circulating PUFA levels

We conducted GCTA-GREML analysis to estimate the amount of phenotypic variance explained by genome-wide gene-FOS interactions. The total additive genetic influence (i.e., SNP heritability) across all 14 PUFA and MUFA traits ranged from 14.2% to 23.9% (s.e. 0.36% and 0.37%) in LA and MUFAs%, respectively (Figure 4, Table S6). The phenotypic variance explained by genome-wide gene-FOS interactions ranged from 0.53% to 1.51% (s.e. 0.38% and 0.39%) in omega-6% to DHA%, respectively. These estimates were statistically significant in 10 out of 14 PUFA traits. The four traits without significant variance explained are total PUFAs, total MUFAs, MUFAs%, and omega-6%. We then estimated the contribution of the top SNP, rs35473591, to the variance explained by gene-FOS interactions in omega-3%. SNP rs35473591 explained a proportion of 5.12 x 10^−4^, estimated to be 4.11% of the genome-wide gene-FOS interaction contributions to omega-3% variance. Gene-FOS interactions make small but non-negligible contributions to the phenotypic variance of plasma PUFA traits.

**Figure 4.**
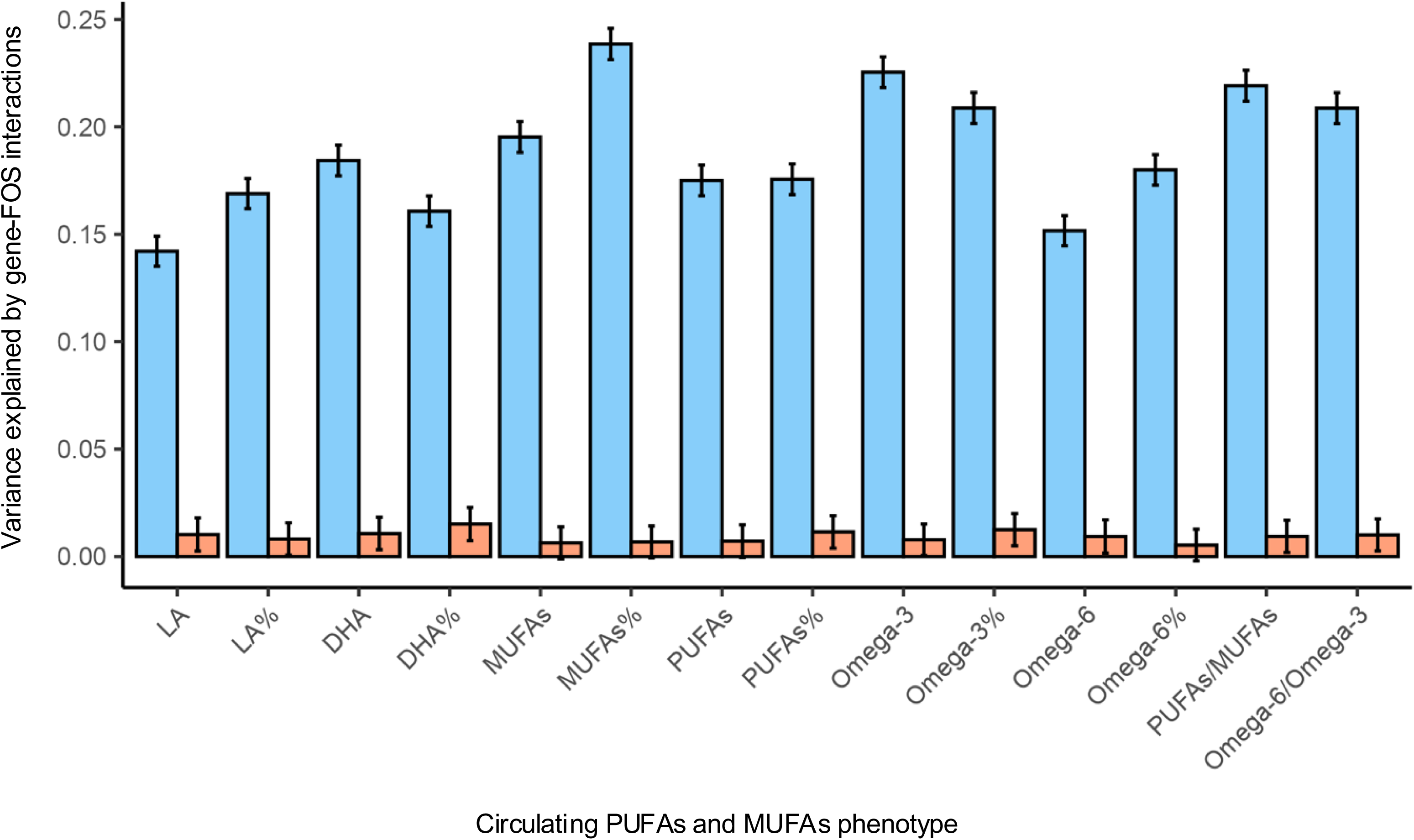
The proportion of phenotypic variance explained by genome-wide gene-FOS interactions in the combined dataset.

## Discussion

In this study, we reported the most extensive GWIS of gene-FOS interactions in 14 PUFAs and MUFAs-related traits. Our goal for this project was to leverage a large cohort of over 200,000 European individuals in the UK Biobank to identify genetic loci that modify the associations between FOS and circulating PUFA traits. We also estimated the contribution of genome-wide interactions with FOS to the phenotypic variance. Across 14 traits, we identified significant gene-FOS interactions for seven traits – five variant and gene-level interactions in omega-3 related traits and the omega-6/omega-3 ratio, and two additional gene-level interactions for PUFAs% and PUFAs/MUFAs ratio.

In both our single-variant and gene-level analyses, we replicated the *FADS1-FADS2* gene cluster identified in previous gene-dietary PUFAs interaction studies.^12,39–41^ These studies investigated other *FADS* SNPs in high LD with our top SNP, rs35473591. These variants do not all have the same effect on PUFAs, so it is essential to investigate each SNP’s effect independently in the different PUFAs.^7^ From our results, the lead *FADS1-* rs35473591 SNP had different effect sizes on omega-3% in the exposure groups. The genetic influence of this SNP was much more substantial in the individuals who do not take fish oil. This shows that FOS lessened the *FADS1-*rs35473591 effect on omega-3%, evidence of interaction. We also saw an overall increase in omega-3-related traits and a reduction in the omega-6 to omega-3 ratio after supplementation, indicating that FOS increases circulating omega-3 levels, as previously established.^4–6,42^ The ratio of dietary omega-6 to omega-3 consumed has drastically shifted from ∼5:1 to 10:1 because of our modern Western diets, and this is reflected in the ratio of plasma omega-6 to omega-3.^43^ It is crucial to regulate this ratio because this shift has been associated with various chronic diseases.^8^

Our results showed that FOS intake had the lowest effect on omega-3% levels in the C/C homozygote genotype group and had the highest effect in the CT/CT group. The degree of omega-3% alteration increased with each copy of the CT allele. This implies that individuals with the CT/CT and C/CT genotypes will benefit more from taking FOS in terms of increasing omega-3 PUFA levels and reducing their omega-6/omega-3 ratio. Individuals who are homozygous of the C major allele at *FADS1-*rs35473591 benefit less, suggesting they need higher doses of FOS to attain the same circulating omega-3 levels as CT carriers. In summary, the *FADS1-*rs35473591 SNP, in the presence of FOS, has an increasing effect on omega-3-related traits, but the degree of this effect is genotype-dependent. We did not find any significant interaction for *FADS* variants in omega-6-related traits. Previous studies have implicated the *FADS* locus in interactions with PUFA supplementation.^13^ A reasonable explanation is that our study focused on omega-3 PUFA supplementation (i.e., FOS), while Sergeant et al.^13^ focused on supplementation of gamma-linolenic acid supplementation, which is an omega-6, 18-carbon PUFA.

Taking only genetic main or marginal effects into account does not give a complete picture of genetic impacts on circulating PUFAs. Accounting for relevant gene-environment interactions revealed patterns that would not be observed in a typical genome-wide association study. Furthermore, revealing gene-environment interactions is needed to estimate how much phenotypic variance is explained by such interactions. This estimation gives a better understanding of complex traits and diseases because they are influenced by a combination of genetic makeup and environmental exposures, and the effects of some genes are environment-dependent. It also helps to explain some of the ‘missing heritability’ in traits. From our results, additive genetic factors alone explain a maximum of 22.5% of the phenotypic variance of omega-3-related plasma levels, while gene-FOS interactions explain a maximum of 1.51%. The contributions of gene-FOS interactions to omega-3 trait variance are small but non-negligible.

Our gene-based analysis revealed the G protein-coupled receptor 12 *(GPR12*) gene as an interacting locus for PUFAs% and the PUFAs to MUFAs ratio. This gene is highly expressed in the central nervous system and encodes the constitutively active G protein-coupled receptor 12 that promotes cyclic AMP production.^44^ GPR12 is an orphan receptor, meaning it is unconfirmed what endogenous ligand it binds to.^44^ Regarding PUFAs, it is unclear how the GPR12 receptor is associated with them. However, the GPR12 receptor is phylogenetically related to cannabinoid receptors, which bind to certain endocannabinoids, including omega-6-derived endocannabinoids.^45,46^ Recent evidence shows that omega-3-derived endocannabinoids also exist.^45,46^ Future research to experimentally establish a connection between PUFAs-derived endocannabinoids and the GPR12 receptor will support our findings.

A significant strength of our study is the large sample with complete dietary, genomic, and NMR PUFA information. We also performed our analyses at both the variant and gene levels and estimated the contributions of genome-wide gene-FOS interactions to the phenotypic variance of circulating PUFA traits, providing the most extensive analysis of gene-FOS interactions. However, our study has several limitations. Firstly, we restricted this study to European-ancestry participants and did not have large enough datasets to replicate these results in other populations. Large enough datasets with the necessary exposure, genomic, and phenotypic information to detect interactions are still uncommon. Secondly, for FOS information, we relied on only one dietary questionnaire administered to participants at one time point. This questionnaire did not give estimates of the frequency of FOS intake, such as daily or weekly. It also did not provide information on the types and compositions of FOS. Hence, we could not account for dosage heterogeneity in our model. Thirdly, other dietary sources of omega-6 and omega-3 PUFAs, such as vegetable oils and fatty fish, can alter circulating PUFA levels.^5,6,42^ Furthermore, FOS status is correlated with other lifestyle and socioeconomic factors.^47^ There is a possibility that the significant interactions we observed in our results were instead driven by other exposures that we did not explore in this study. However, our study had enough statistical power and was rigorous enough to replicate the *FADS* locus, giving credibility to our findings.

## Conclusion

Our study revealed a novel locus that significantly alters the effect of FOS on PUFAs-related traits. Taking genetic information into account when making nutritional recommendations is the hallmark of precision nutrition, which aims to move away from a one-size-fits-all dietary recommendation model. Variants that influence the effects of dietary PUFAs on plasma PUFAs will help inform diet-based disease prevention and treatment strategies. Additionally, accounting for gene-FOS interactions will aid in developing more accurate polygenic scores for PUFAs-related traits or PUFAs-associated diseases. Lastly, our study also brings to light a potentially unexplored association between the GPR12 gene and PUFAs. A future research direction to experimentally validate this association may highlight PUFA supplementation as a potential treatment for GPR12-related diseases, such as schizophrenia.^48^

## Data Availability

Data processed in this study was made available through the UK Biobank and accessed via an approved application. Data are not publicly available and can be accessed by applying through https://www.ukbiobank.ac.uk/enable-your-research/register. Summary statistics will be available in the GWAS Catalog (https://www.ebi.ac.uk/gwas/) after publication. Scripts used to process the data, perform analysis, and interpret results are available on GitHub (https://github.com/adannasusan/PUFA-GxE).

## Description of Supplemental Information

Supplemental information includes five figures and six tables.

## Acknowledgments

We would like to acknowledge and express gratitude to the UK Biobank participants and administrative staff. Research reported in this publication was supported by the National Institute of General Medical Sciences of the National Institute of Health under the award number R35GM143060 (KY). The content is solely the authors’ responsibility and does not necessarily represent the official views of the National Institutes of Health.

