## Supplementary material for "A multi-level gene-diet interaction analysis of fish oil supplementation and 14 circulating polyunsaturated fatty acids-related traits identifies the *FADS* and *GRP12* loci": Fig S1-S5 Table S1

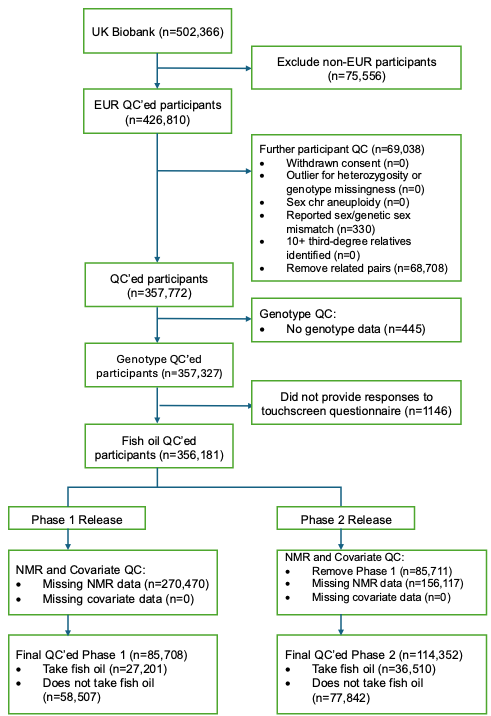

Figure S1. Participant flowchart

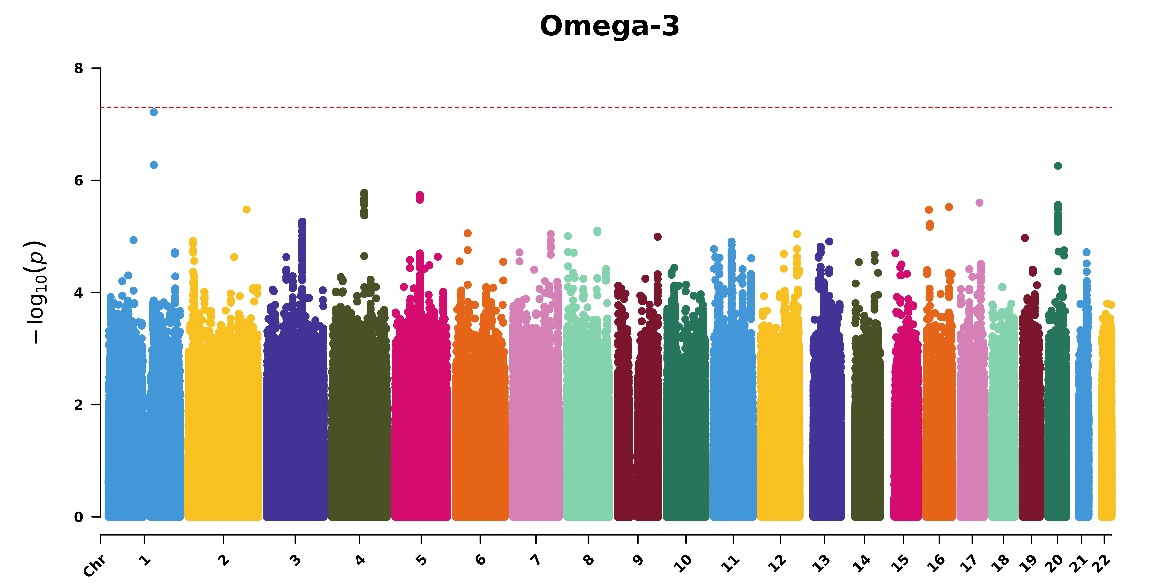

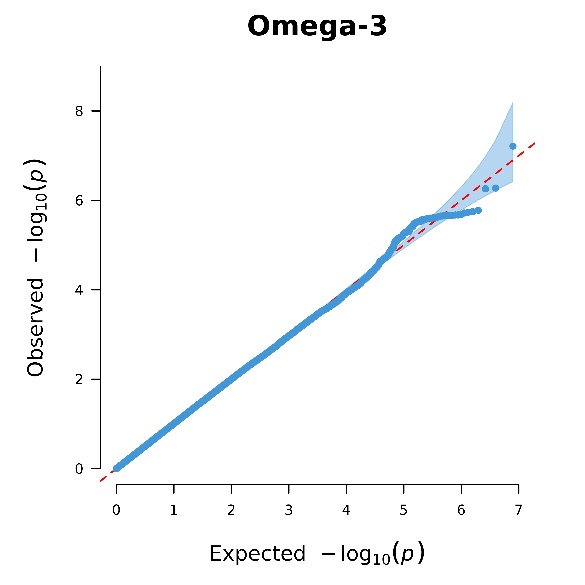

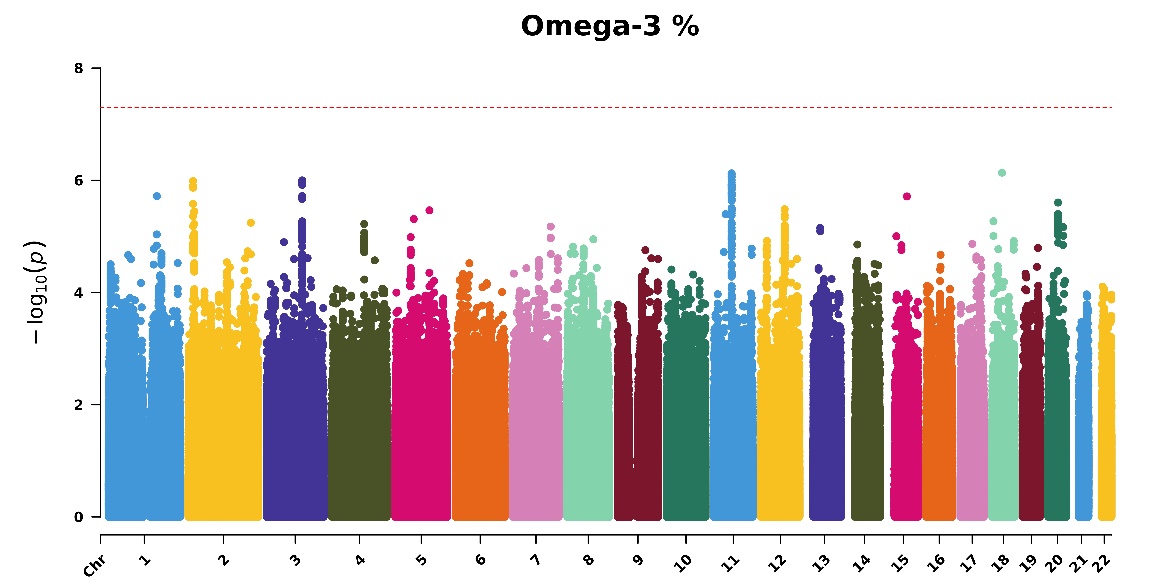

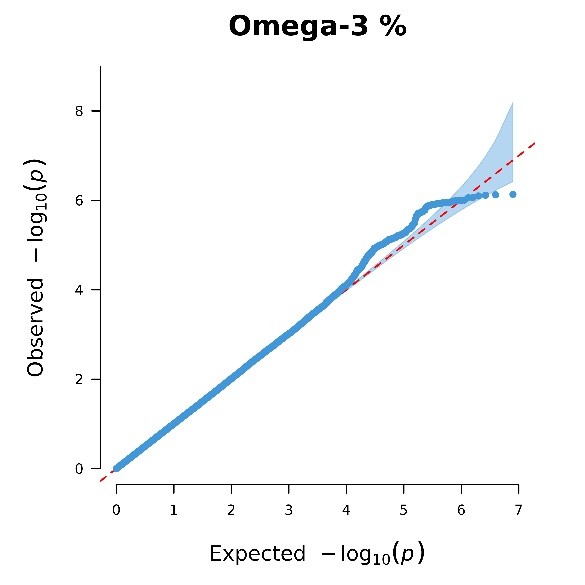

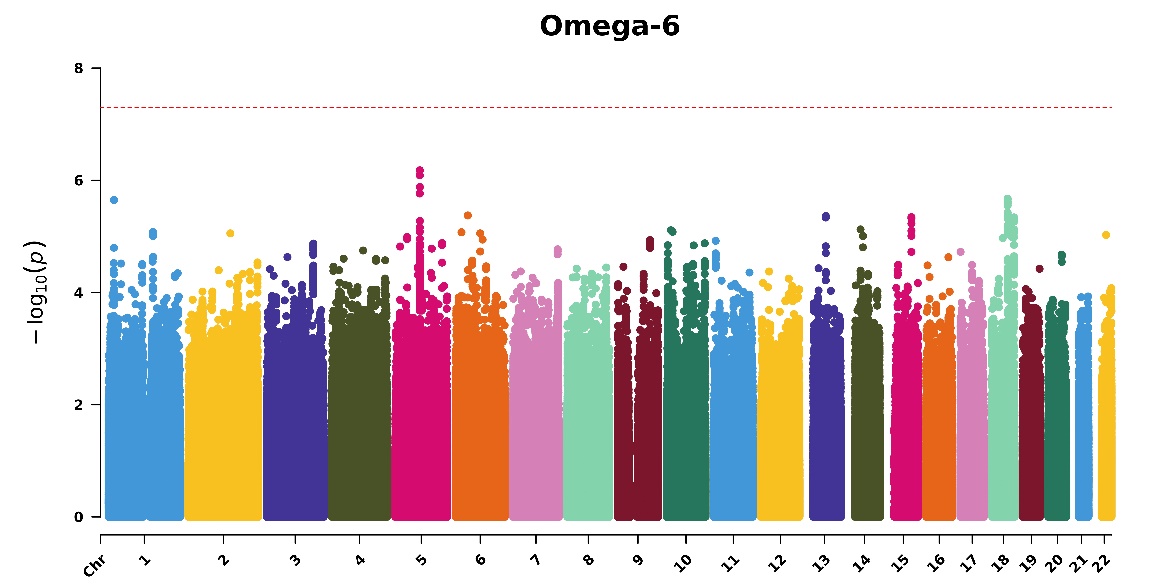

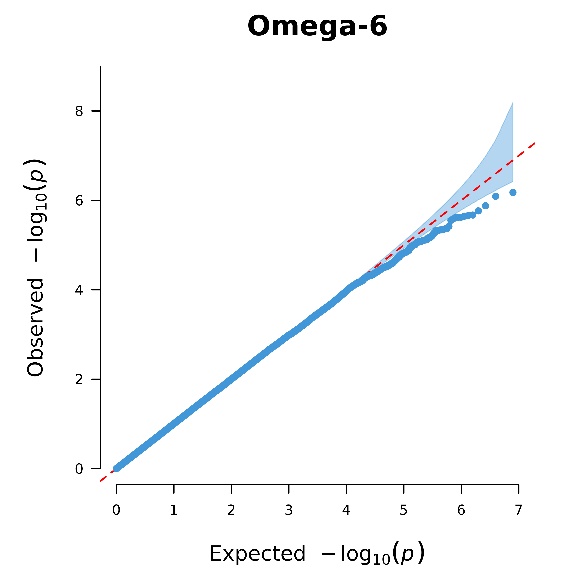

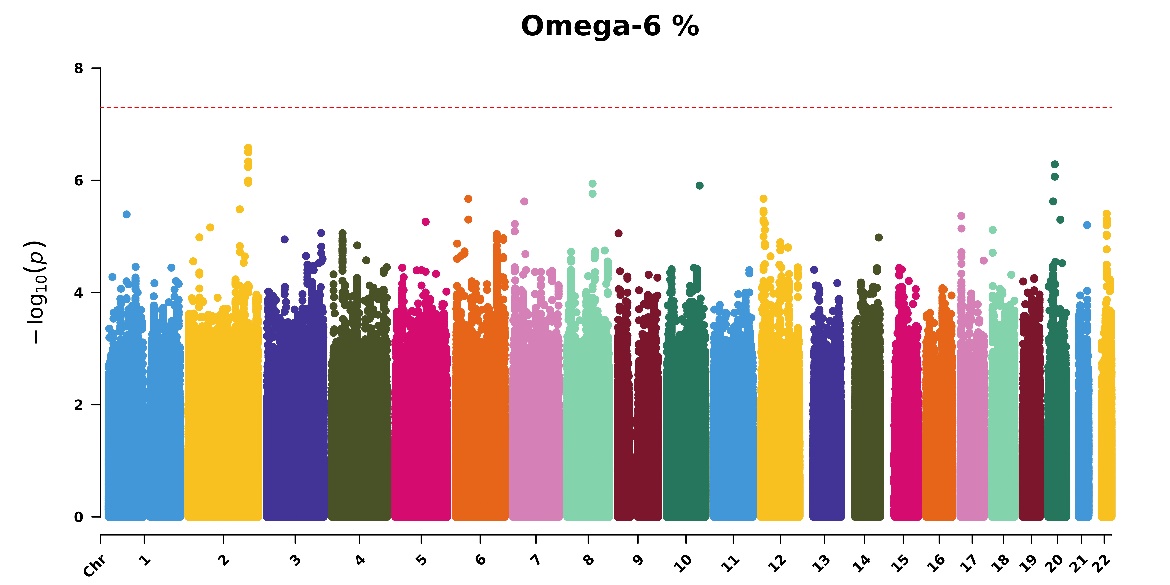

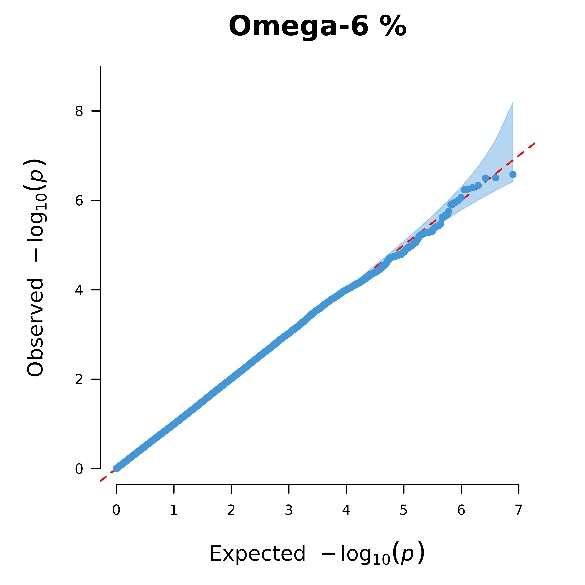

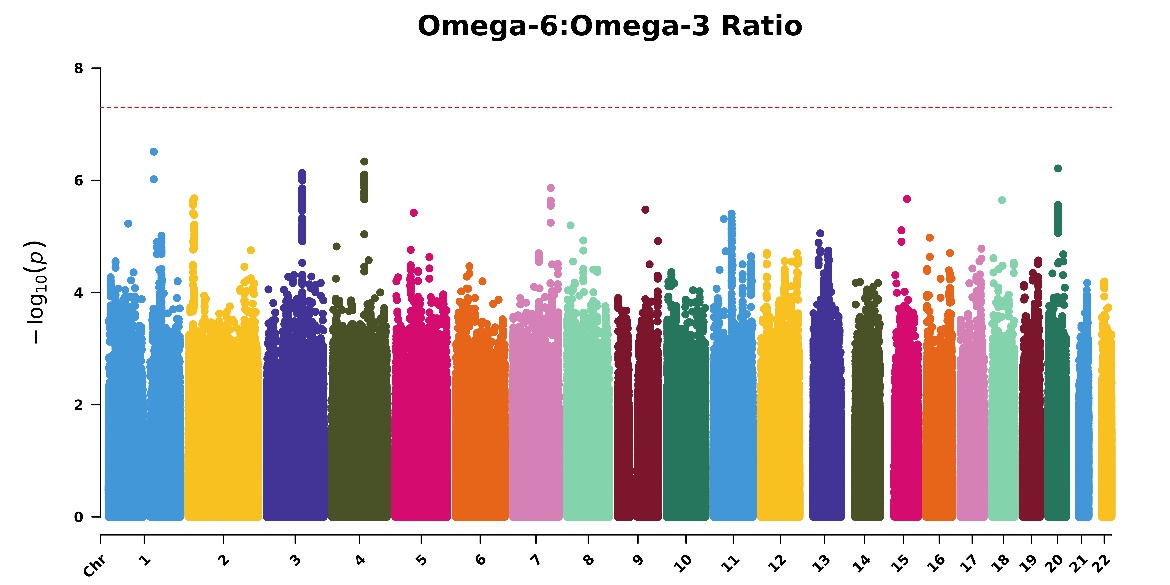

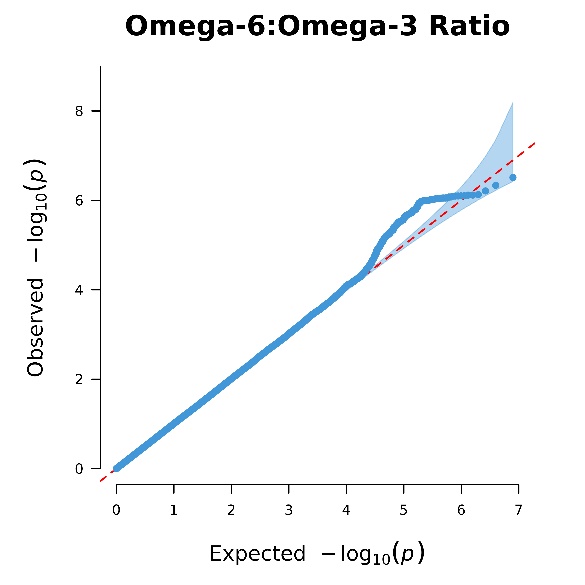

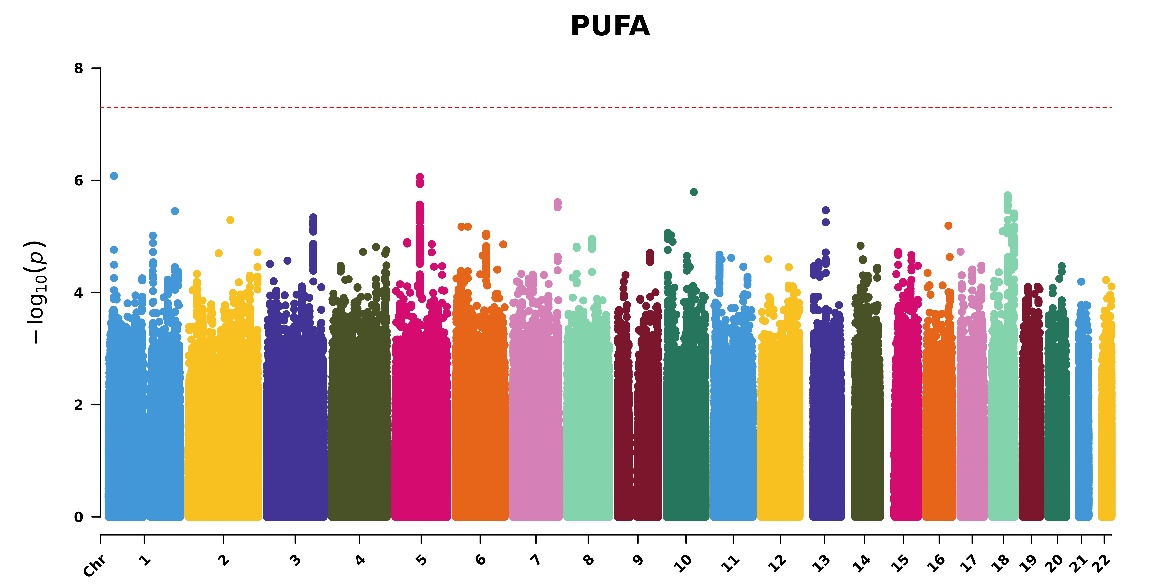

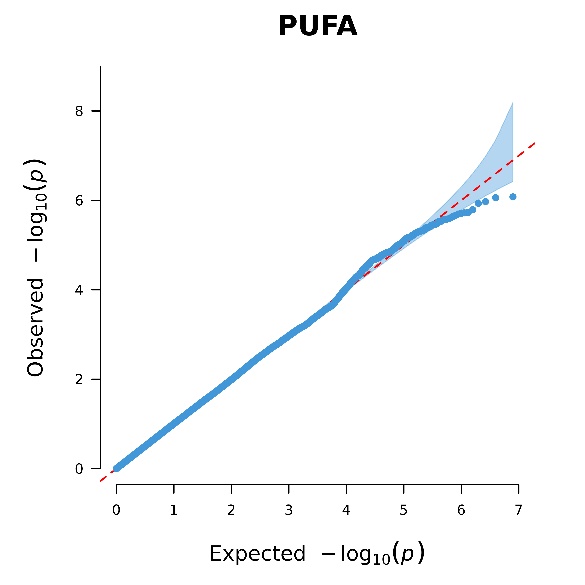

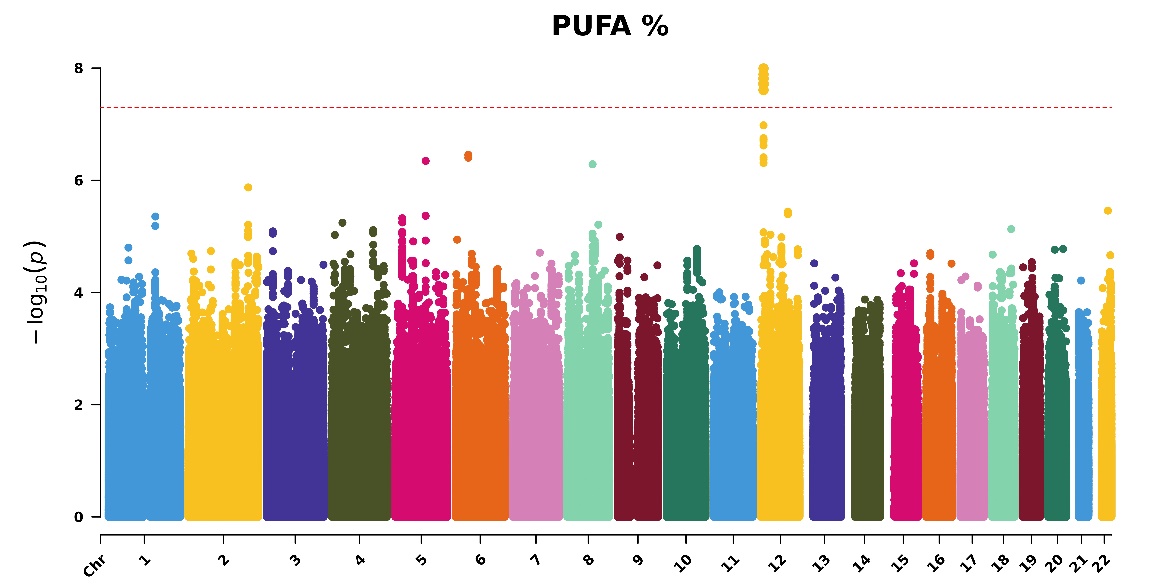

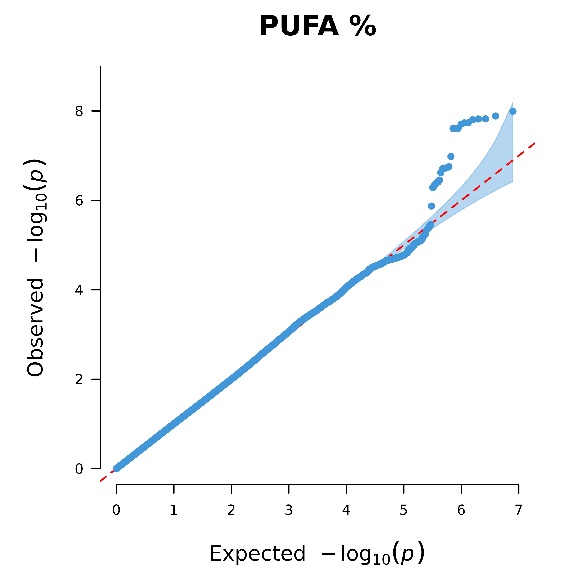

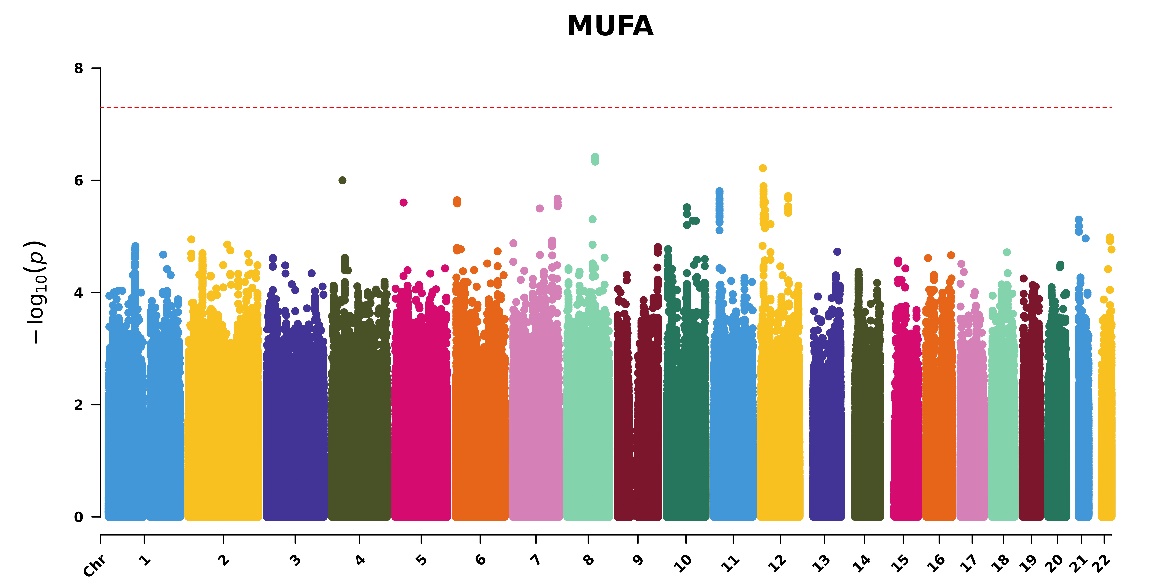

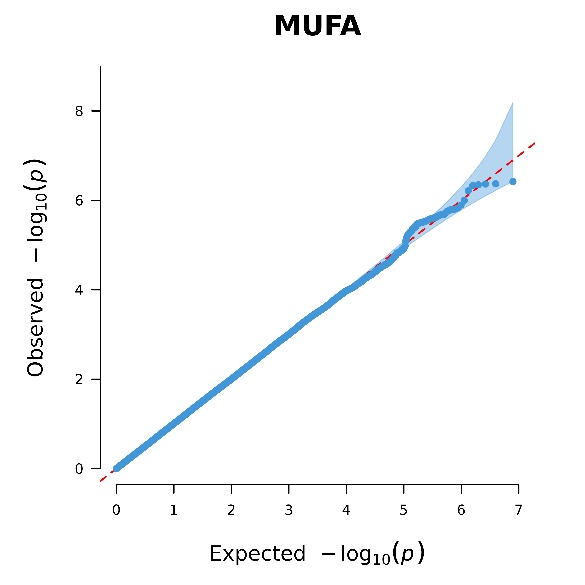

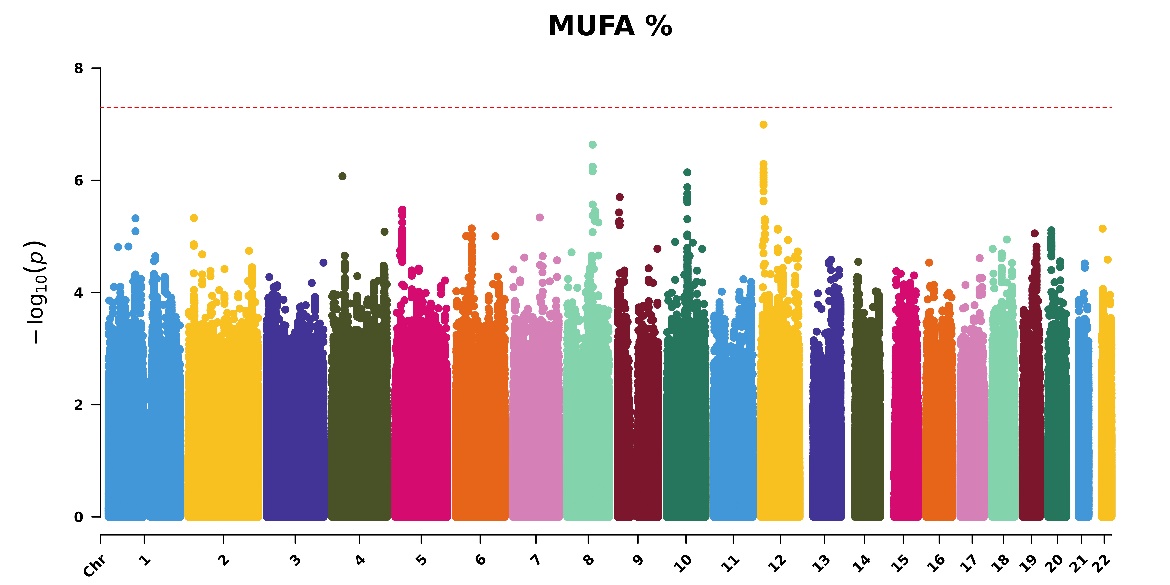

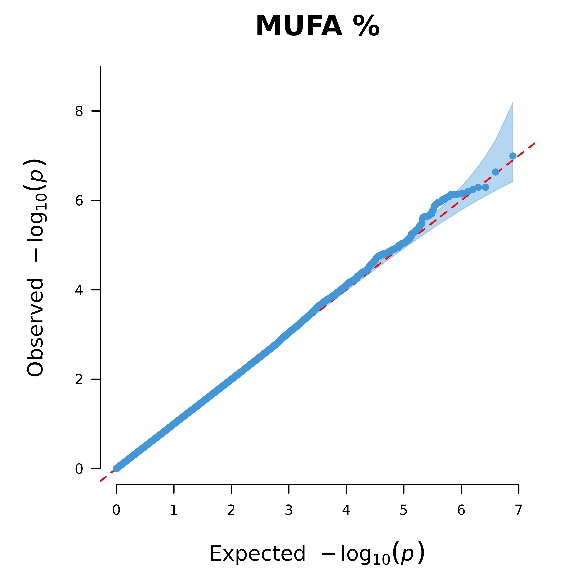

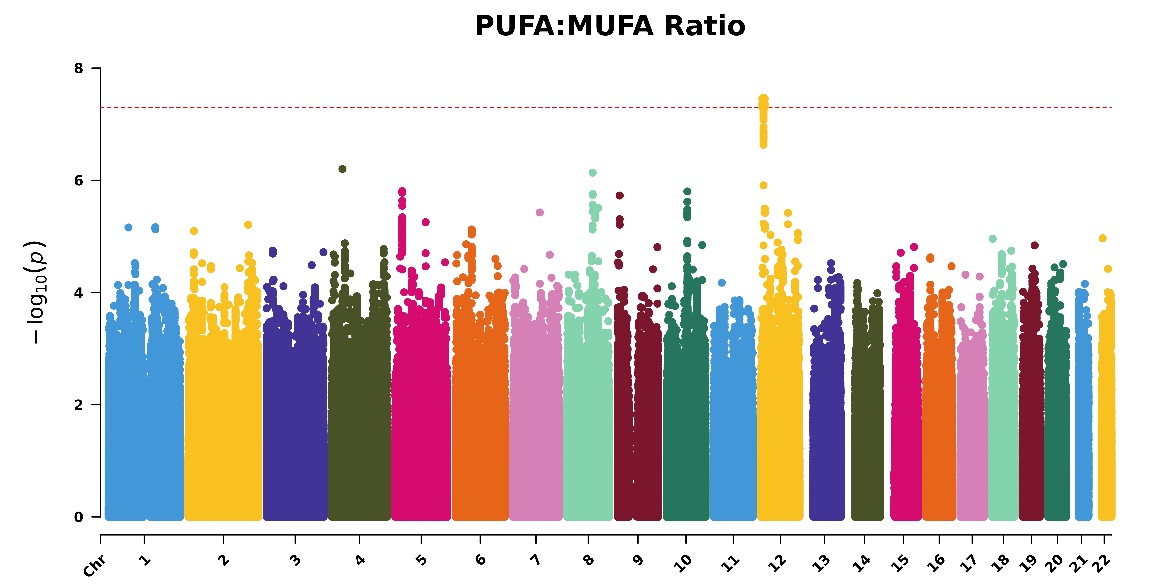

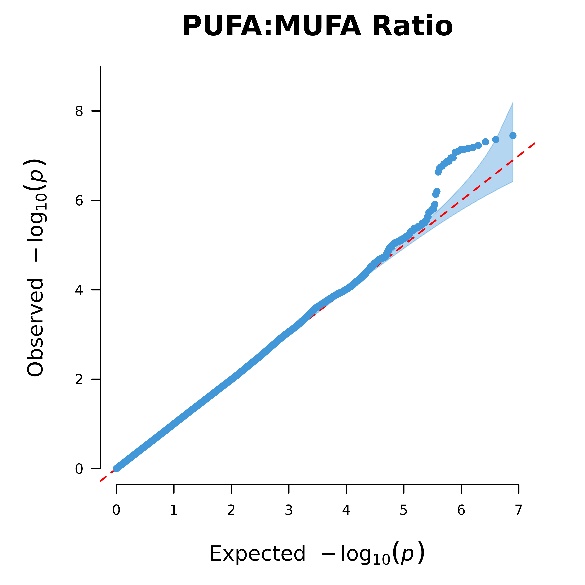

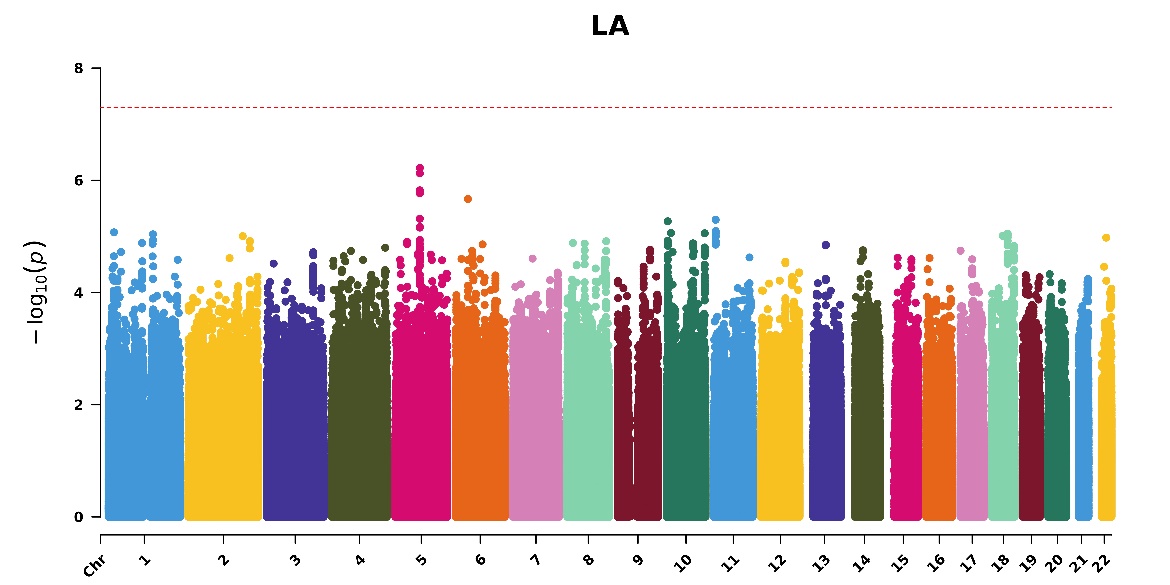

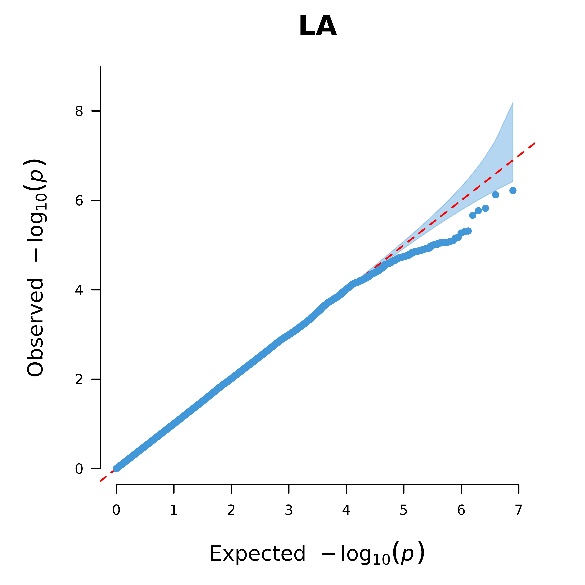

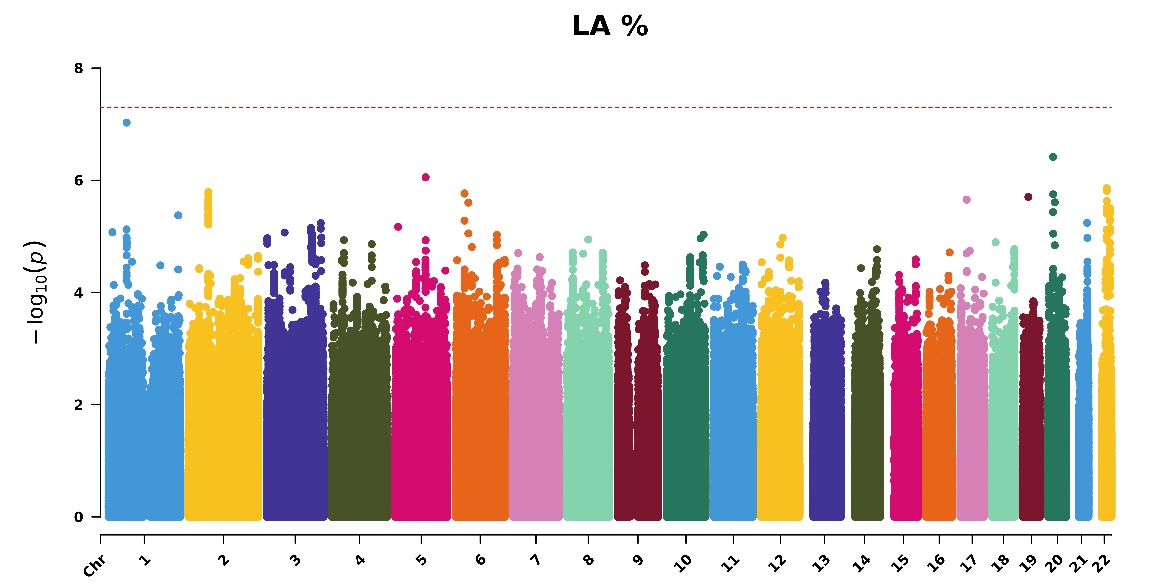

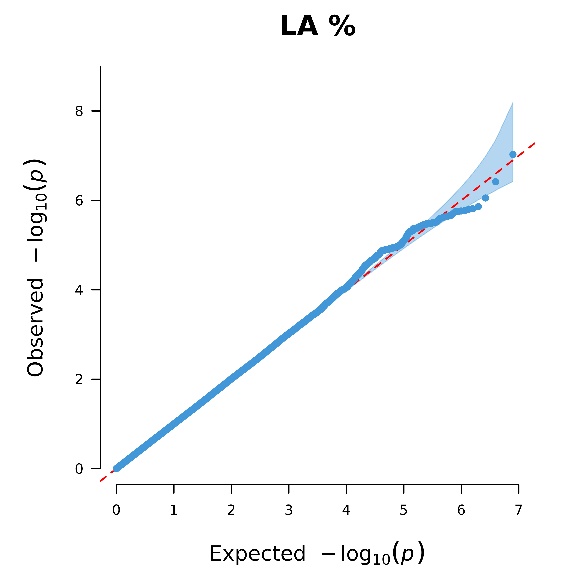

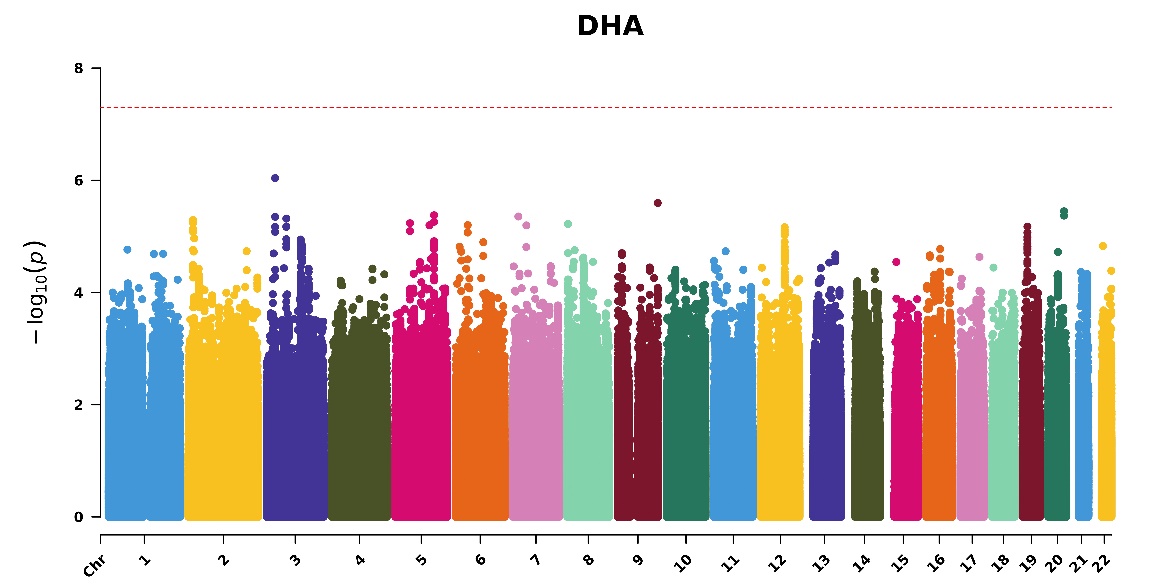

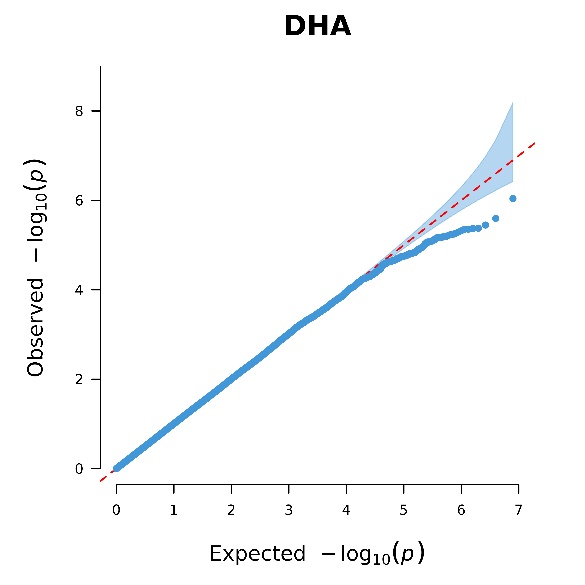

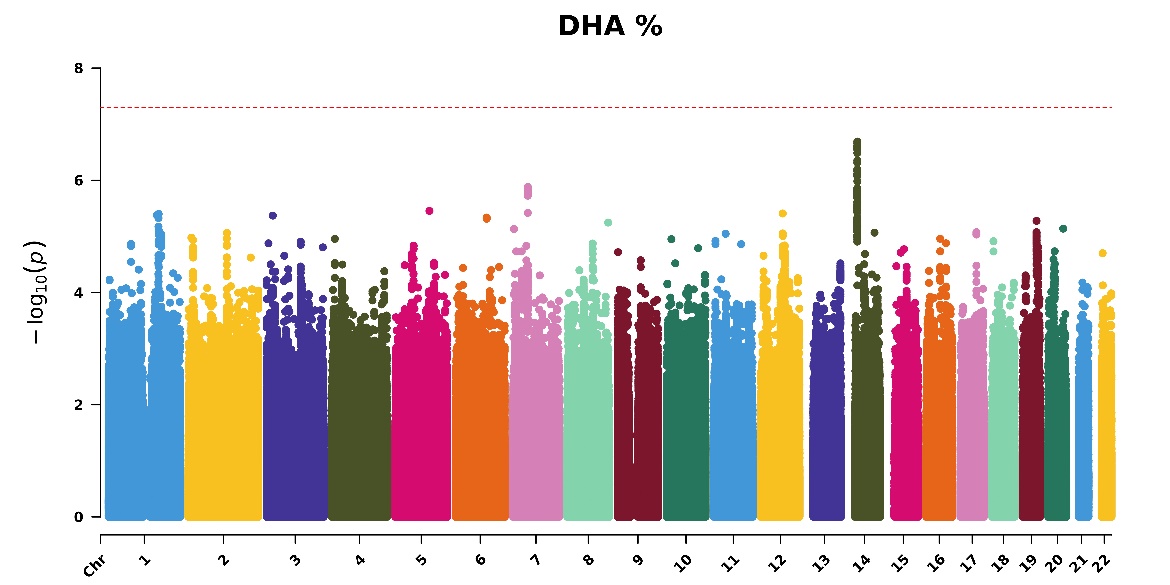

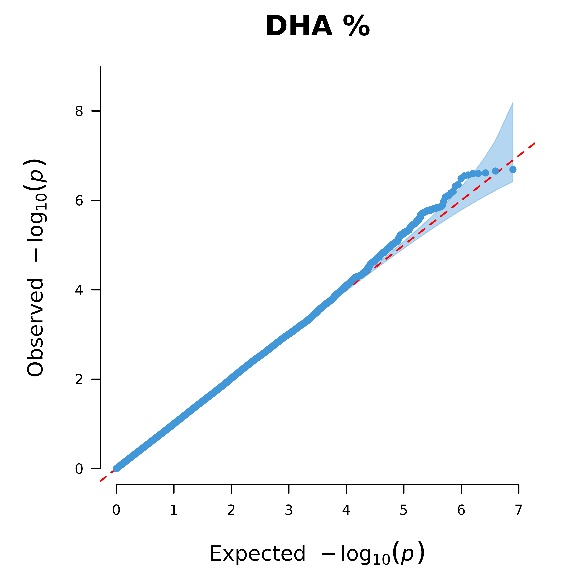

### Figure S2. Manhattan and QQ plots of *p*-values for gene-FOS interactions in 14 PUFAs and MUFAs-related phenotypes for 85,708 participants in the Phase One dataset

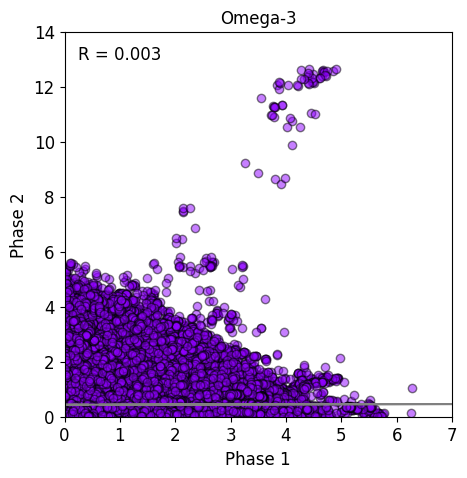

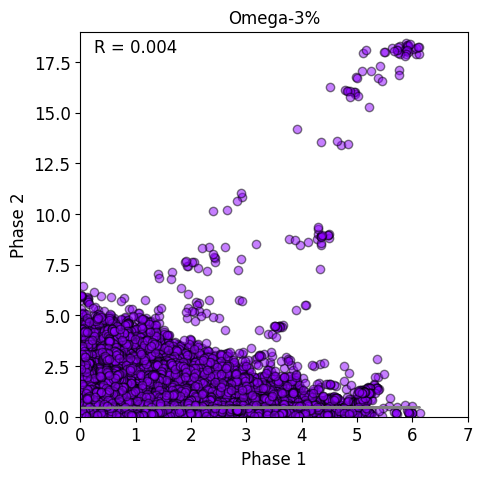

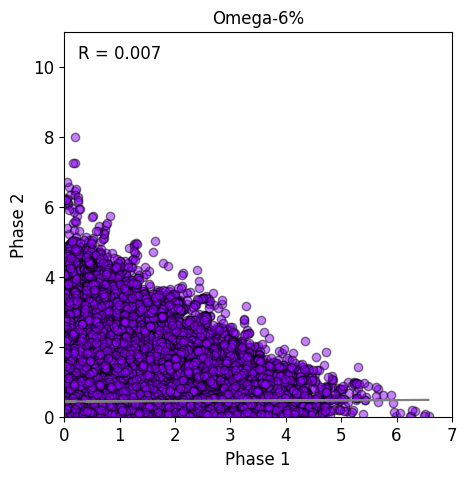

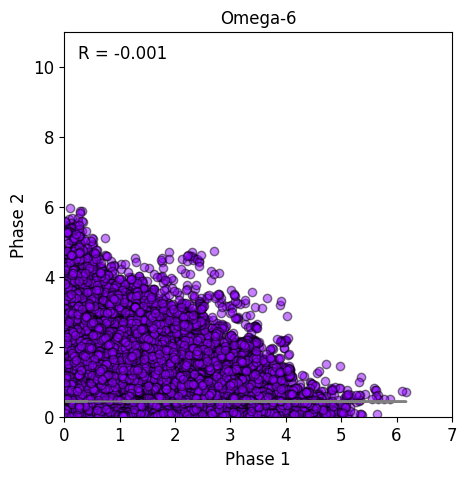

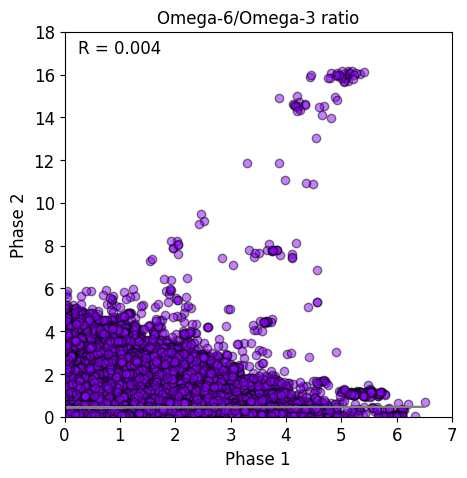

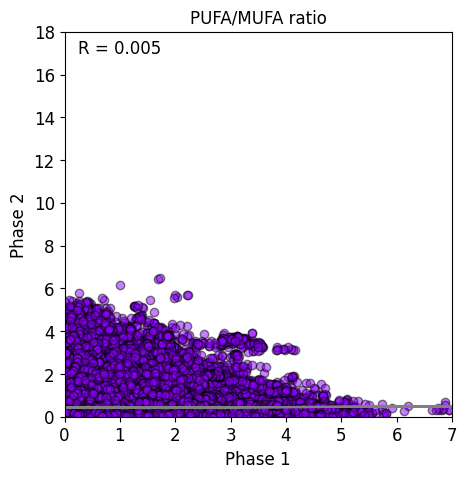

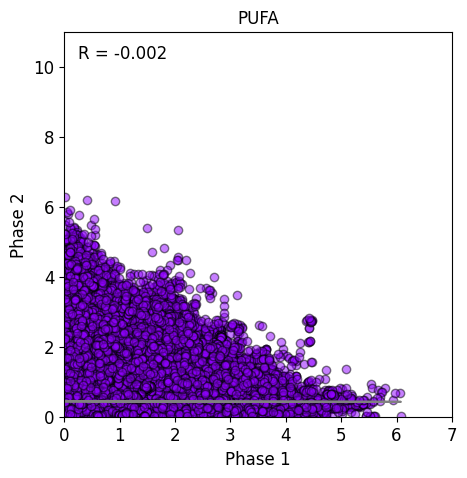

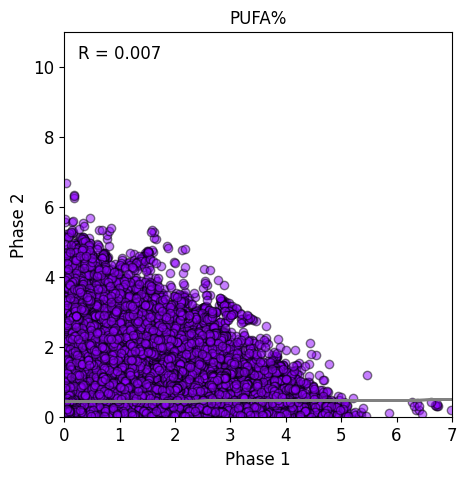

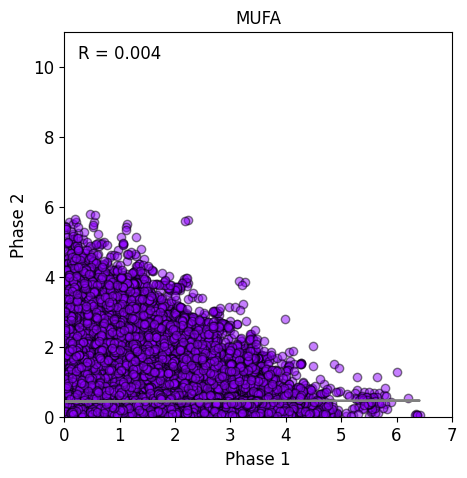

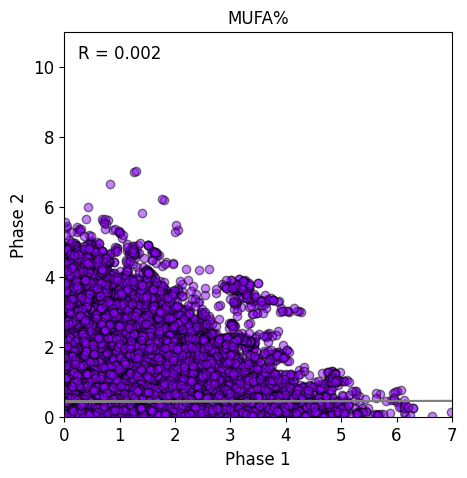

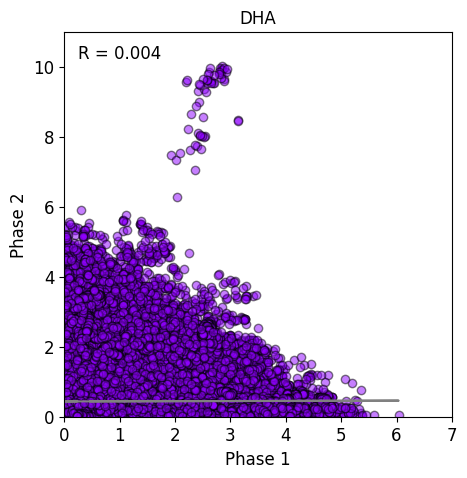

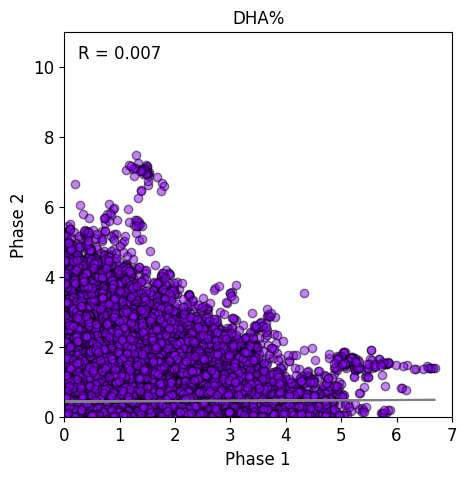

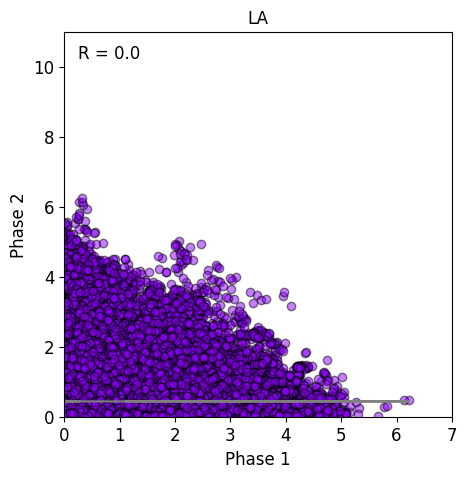

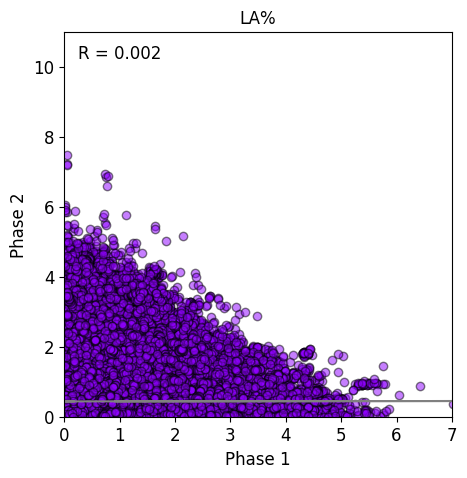

### Figure S3. Scatter plots of *p-*values, across the genome, of gene-FOS interactions across 14 PUFAs and MUFAs-related phenotypes in the Phases One and Two datasets

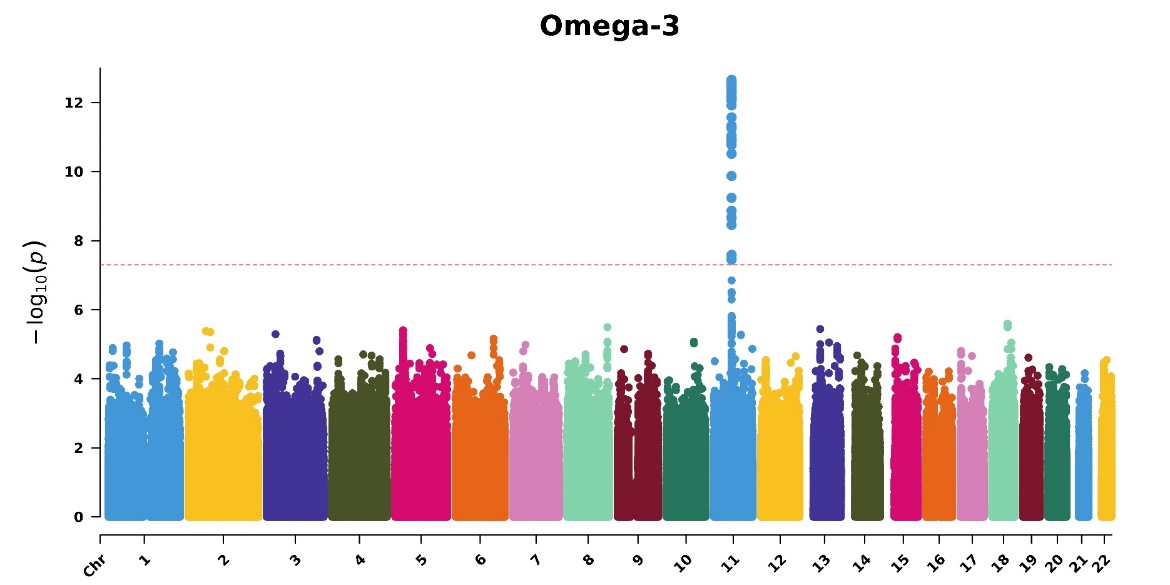

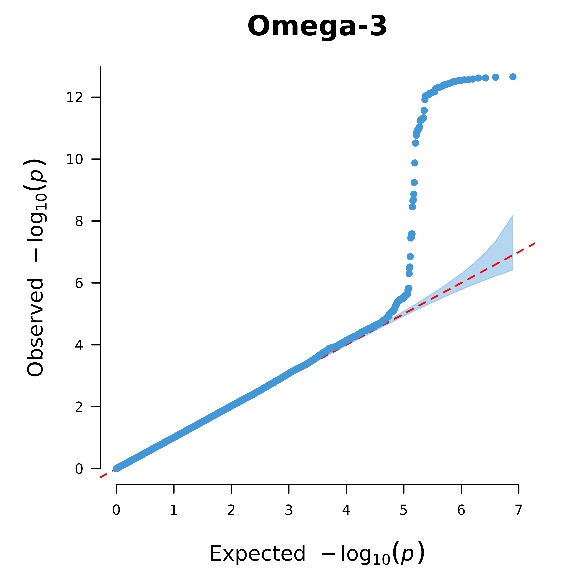

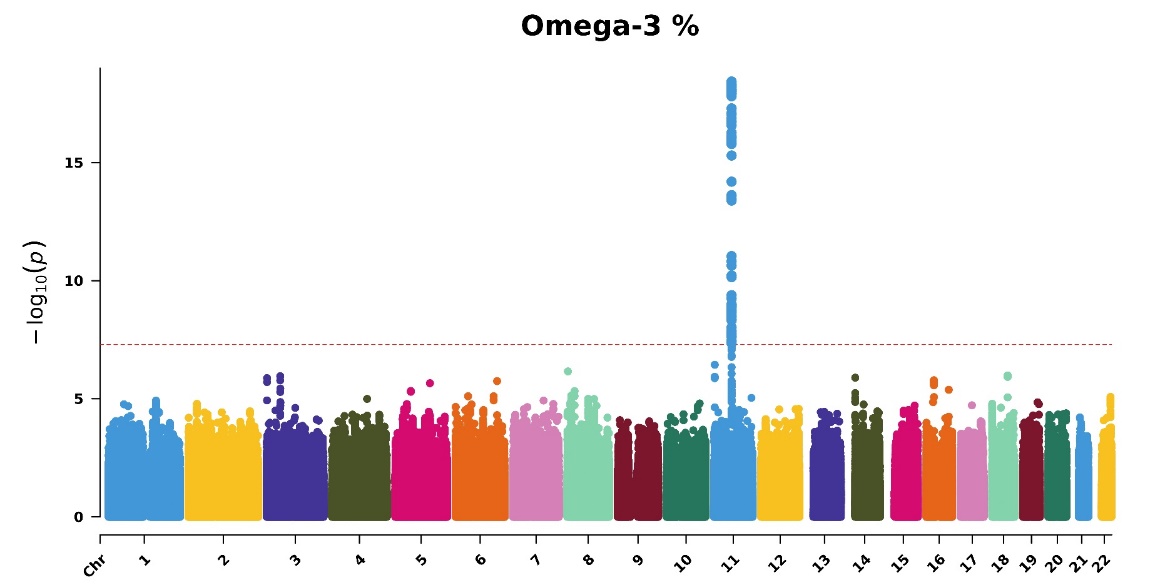

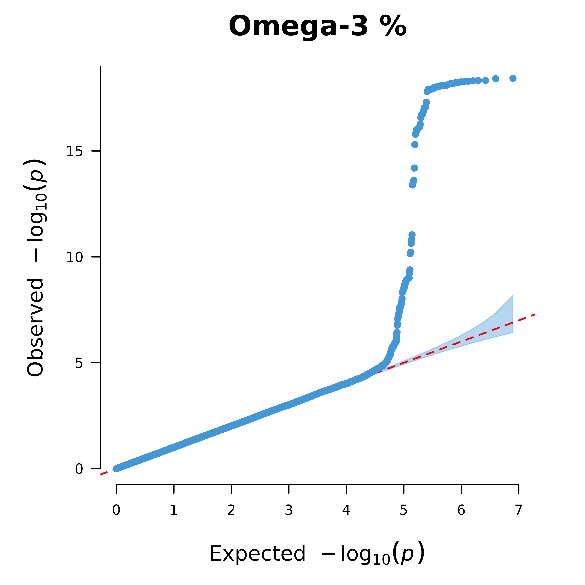

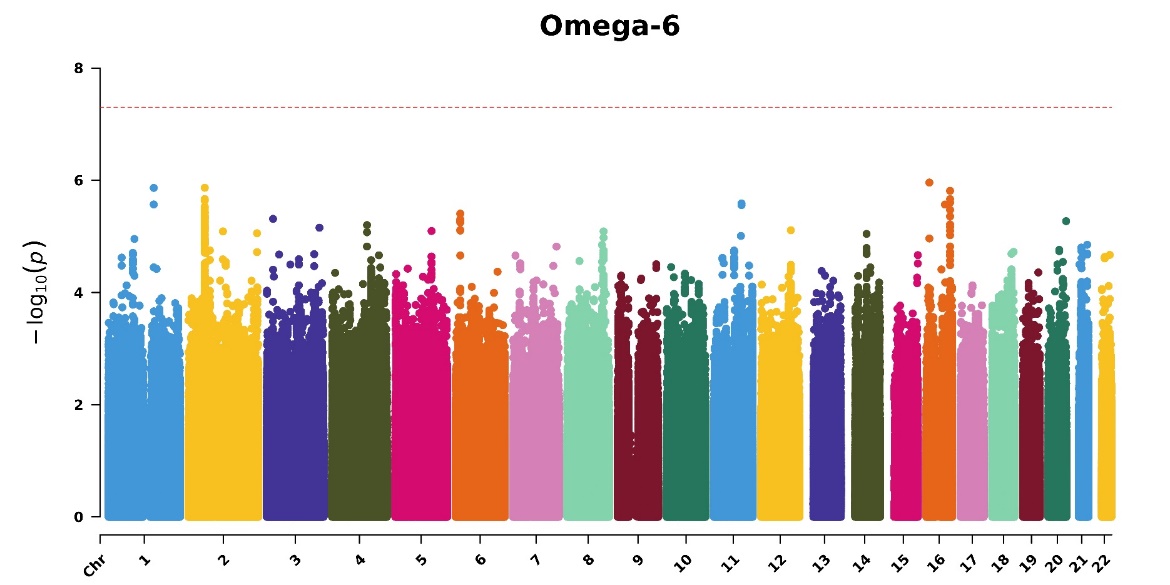

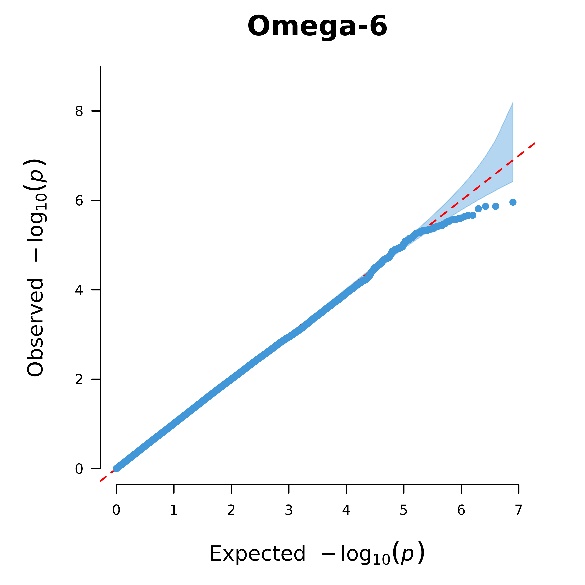

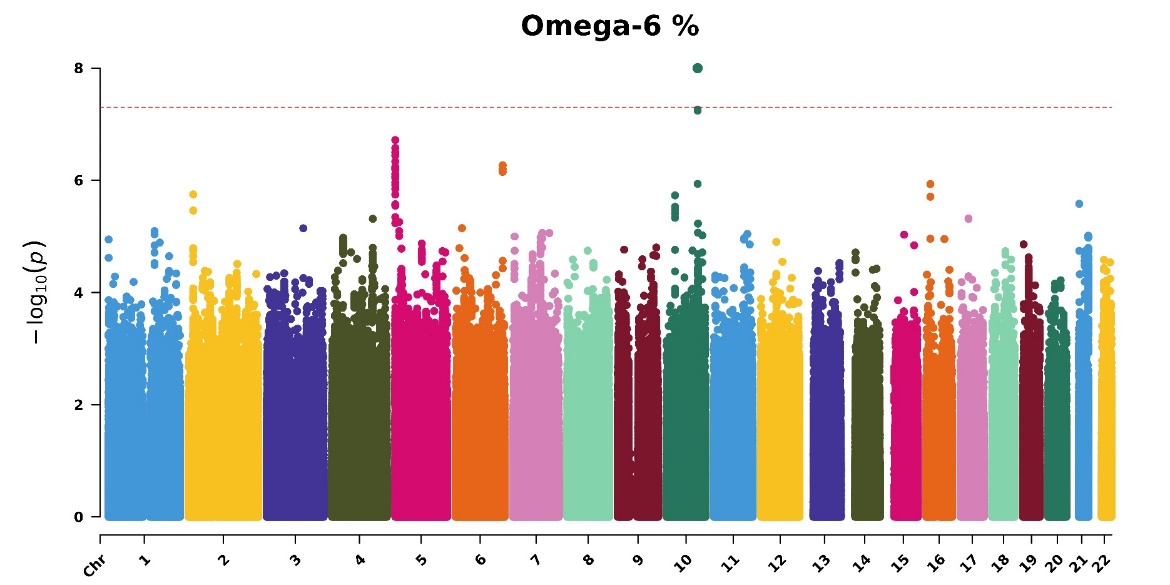

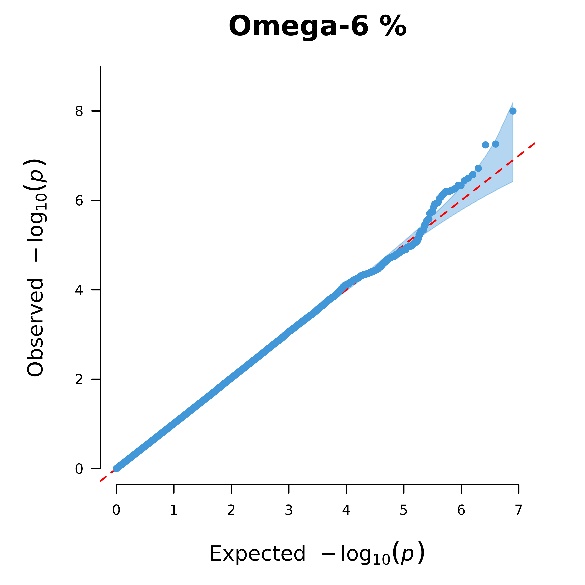

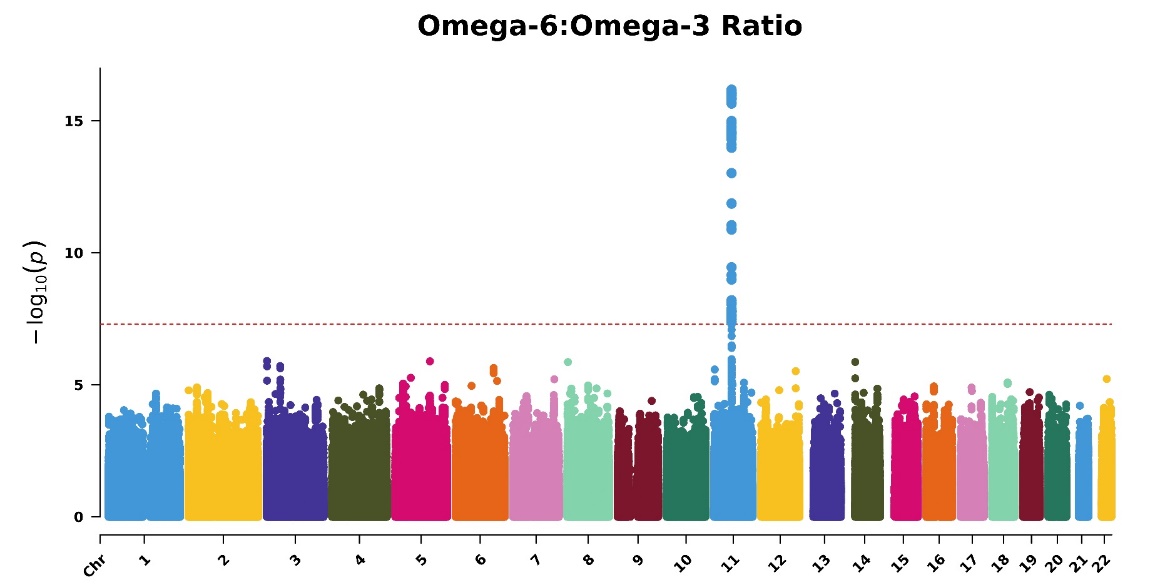

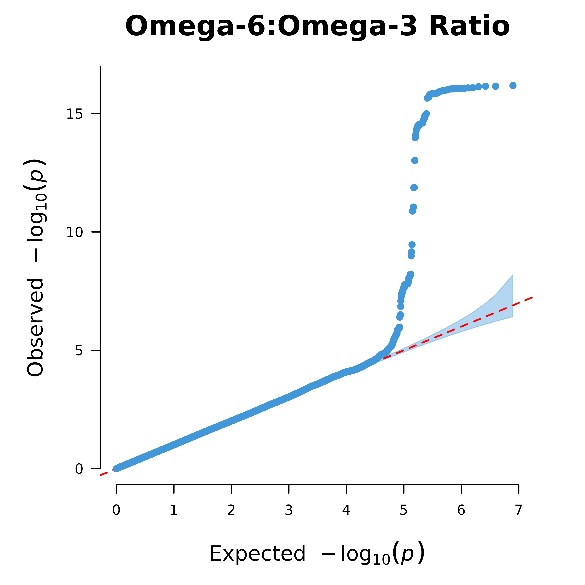

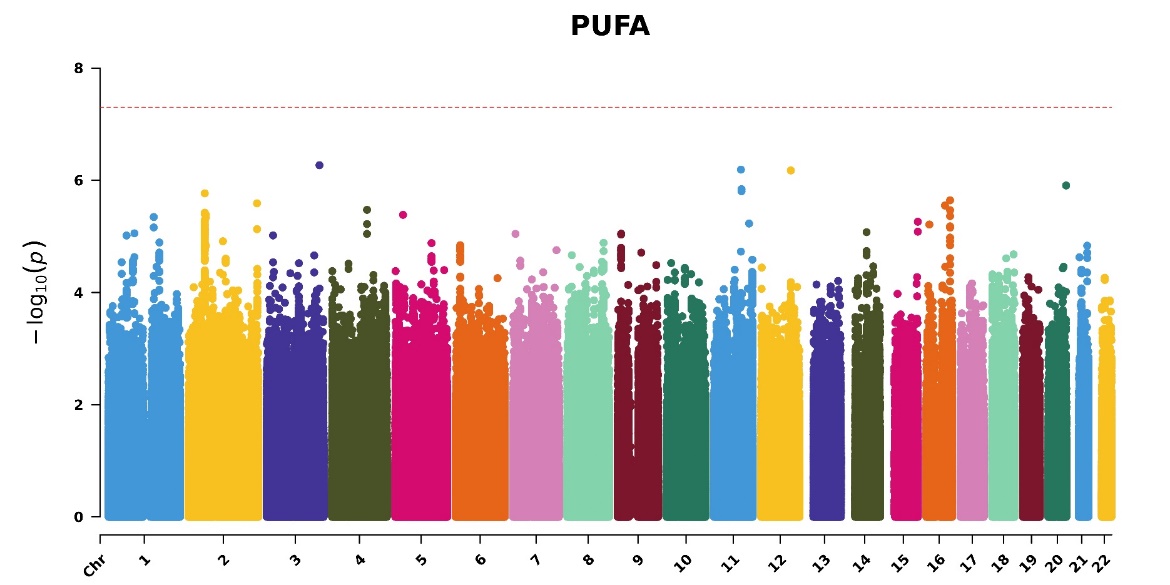

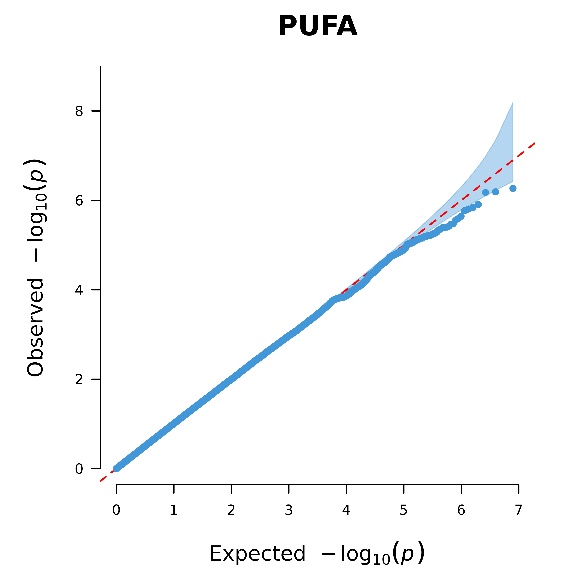

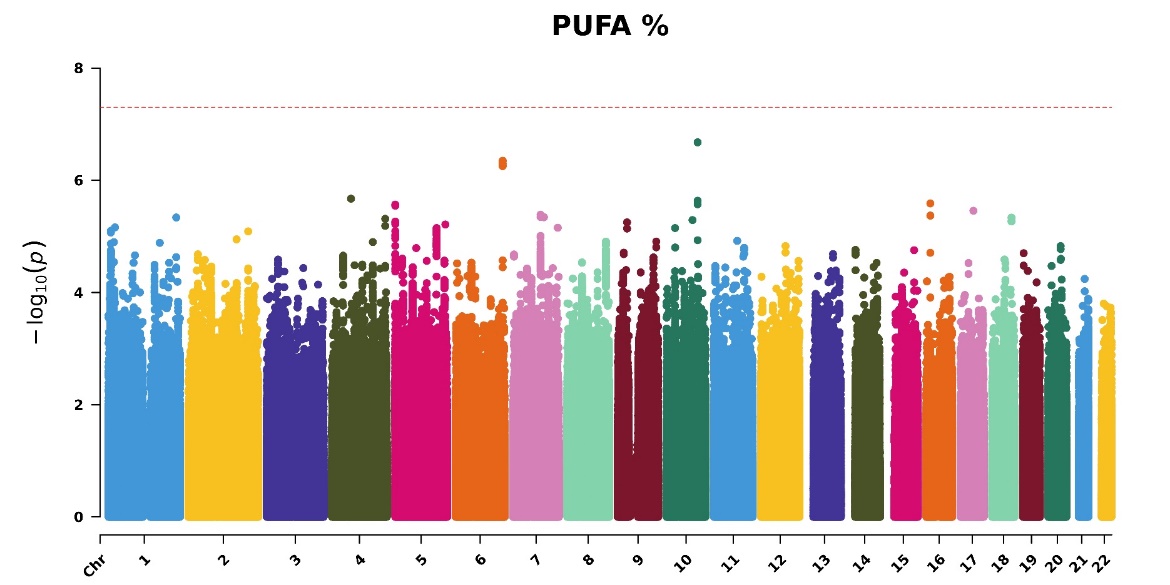

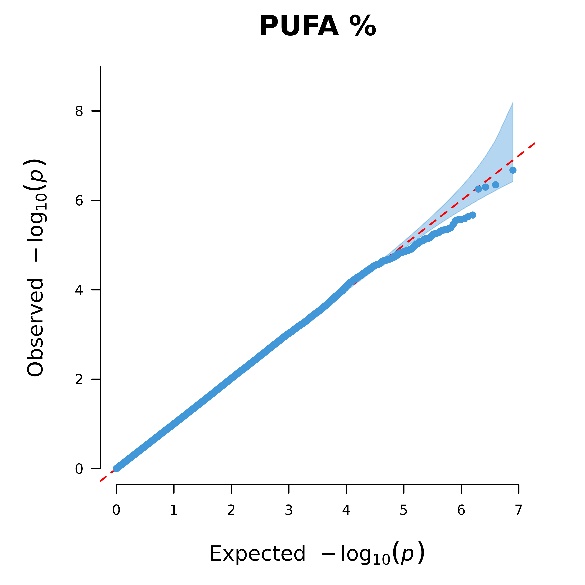

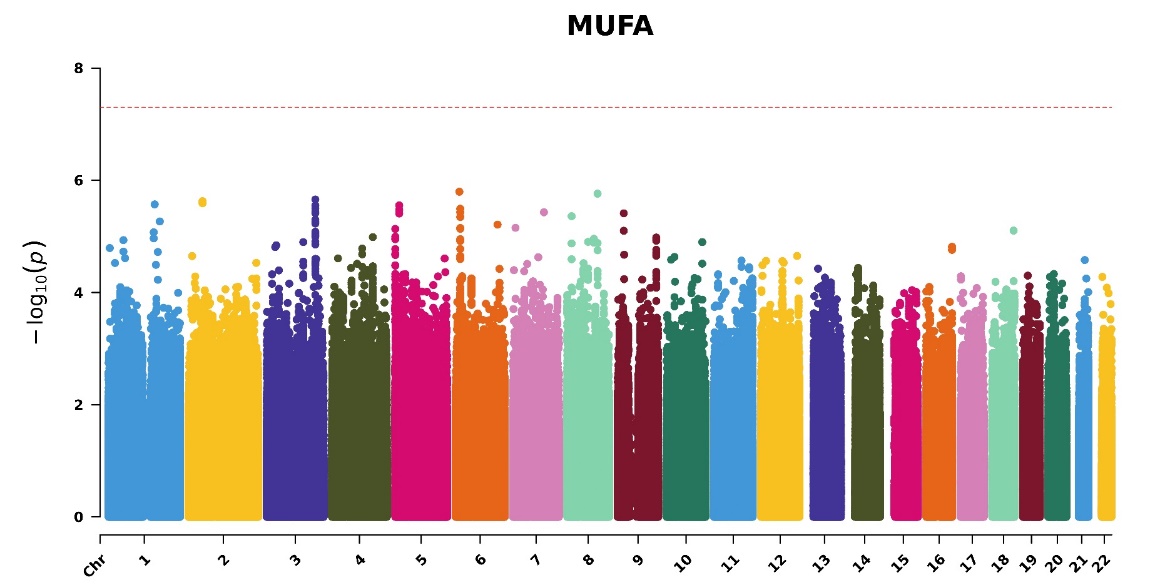

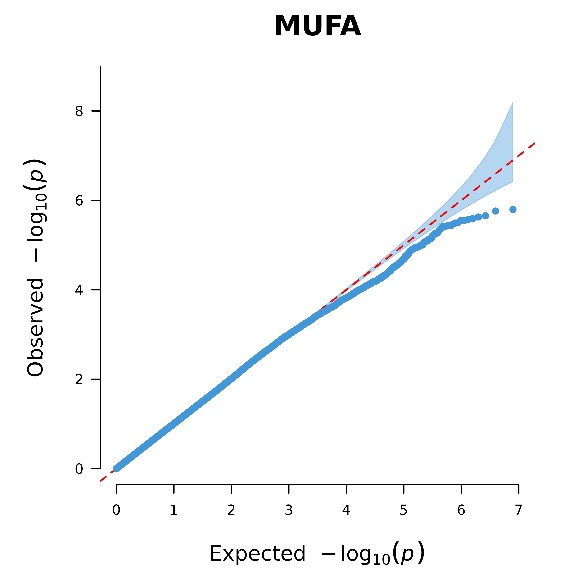

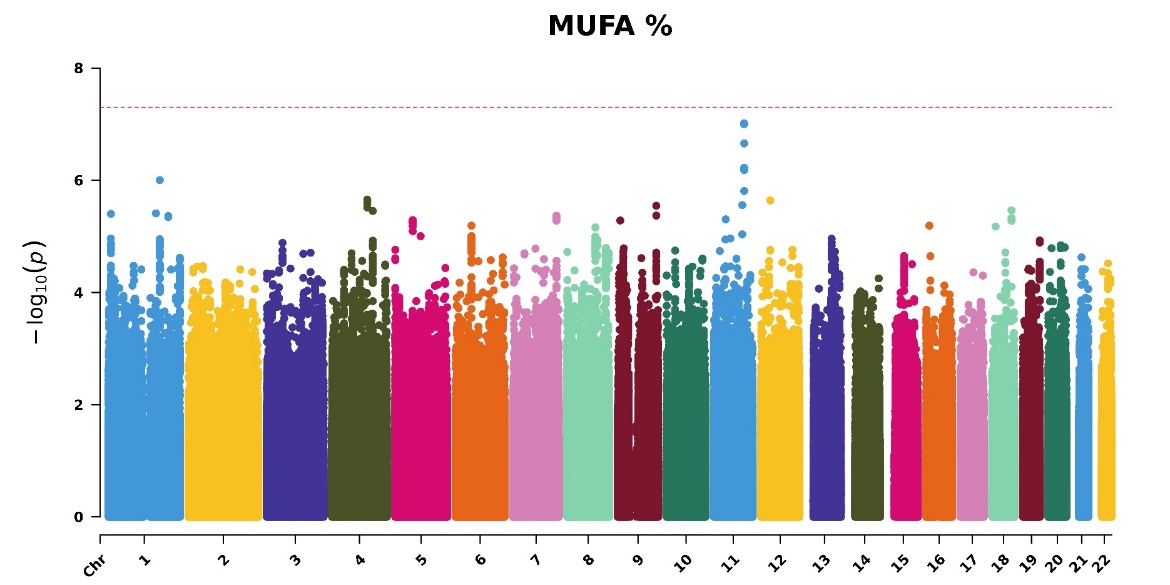

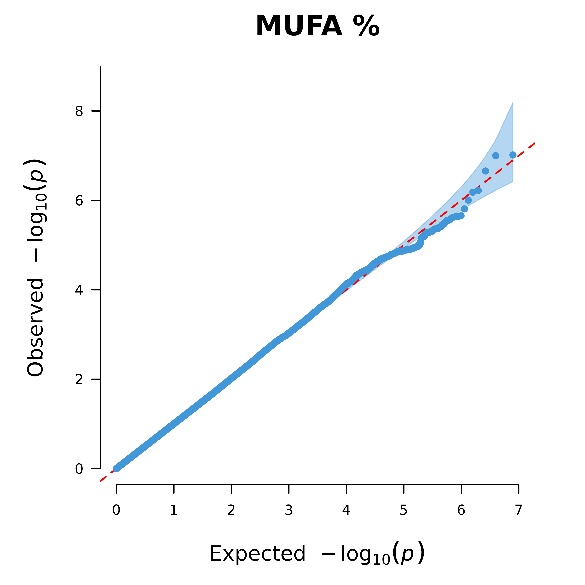

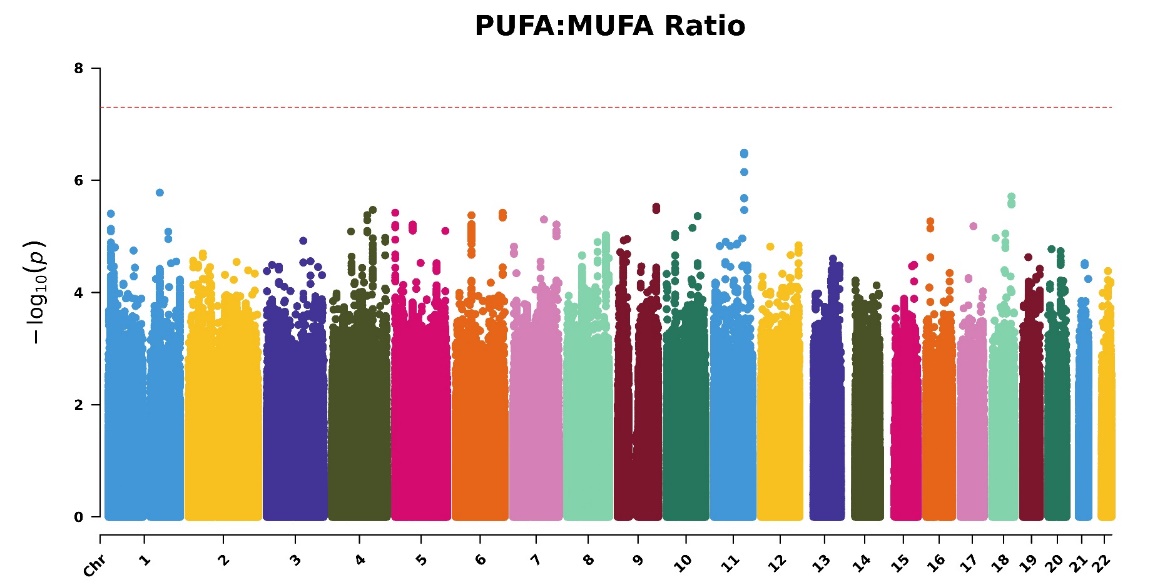

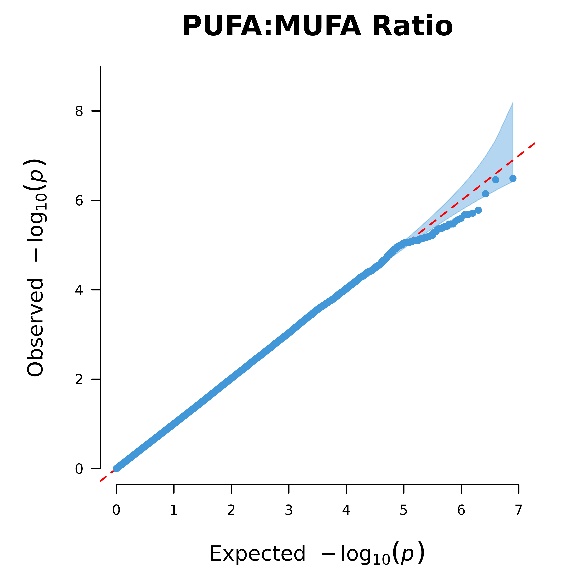

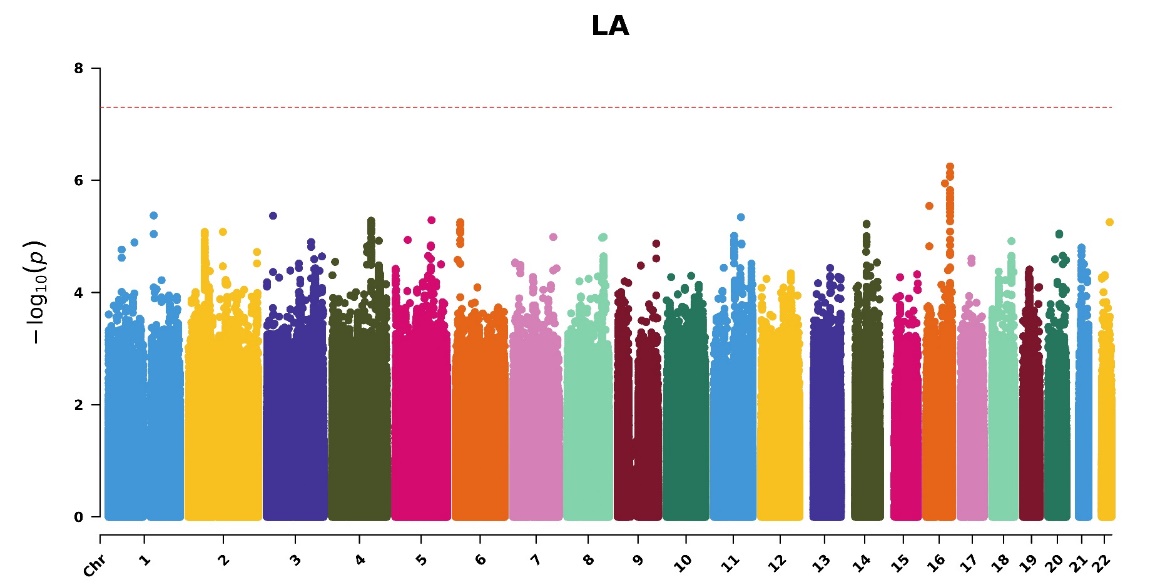

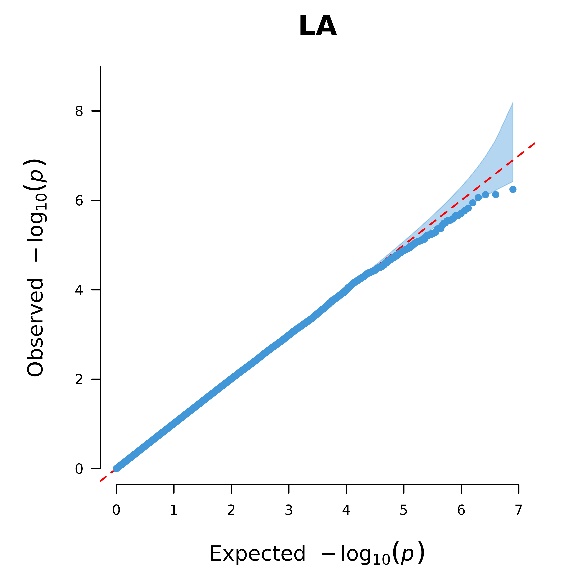

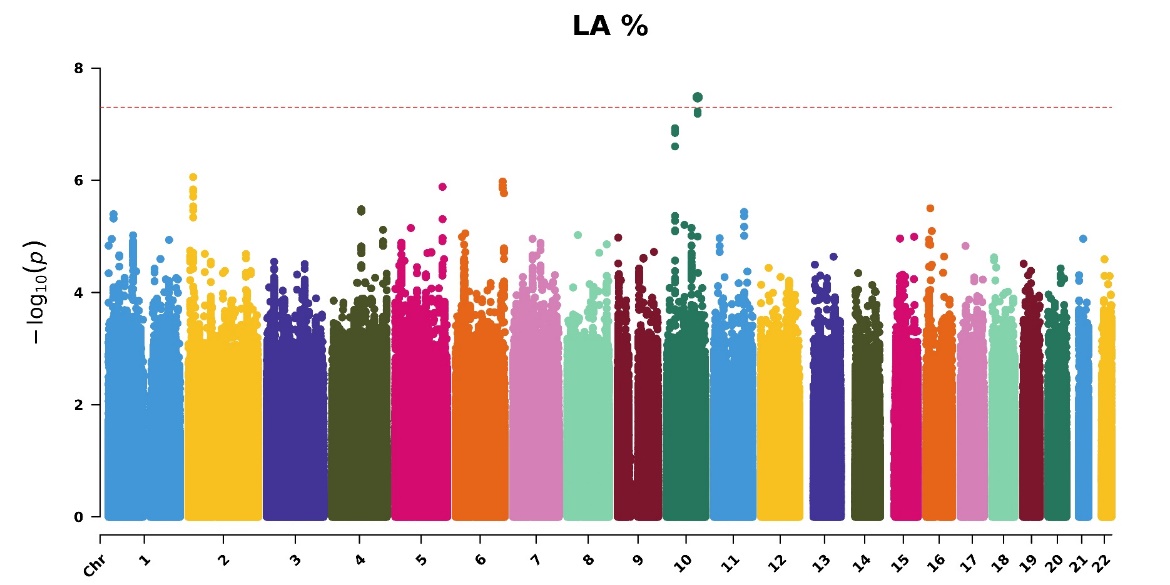

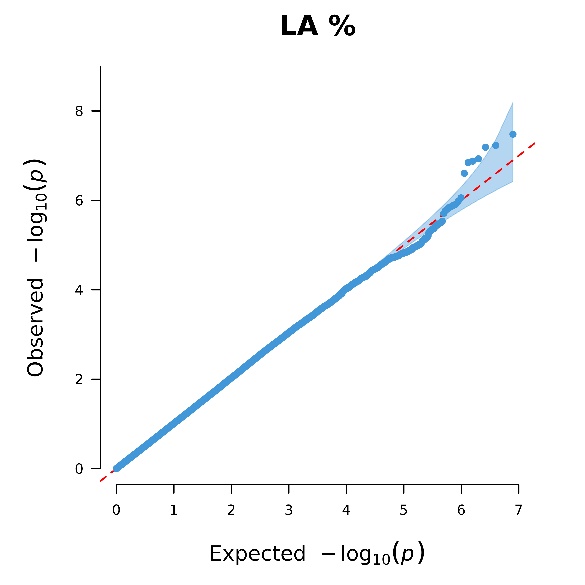

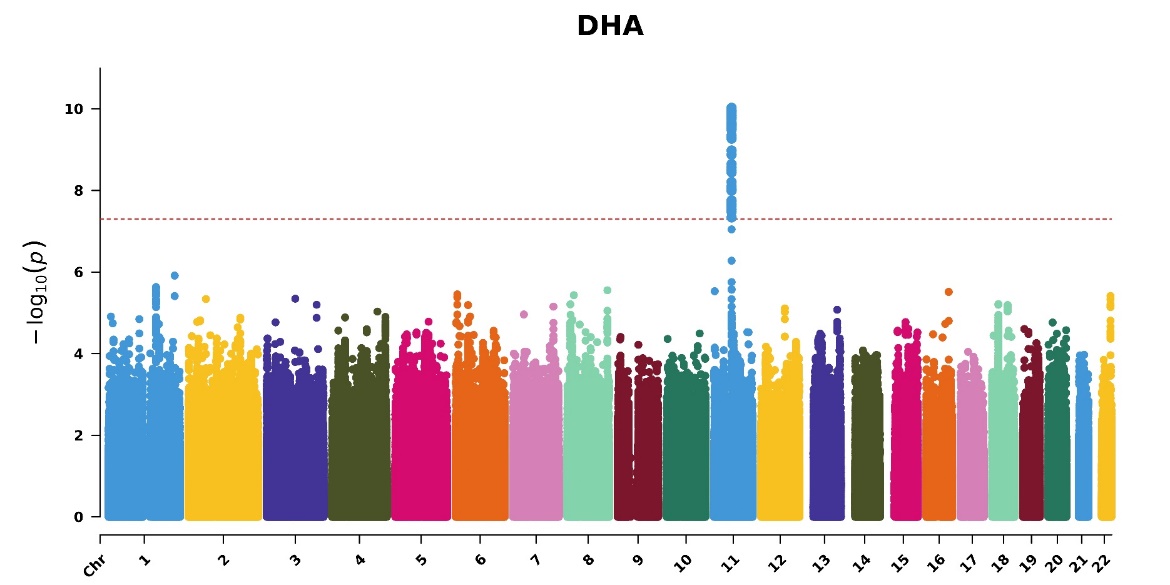

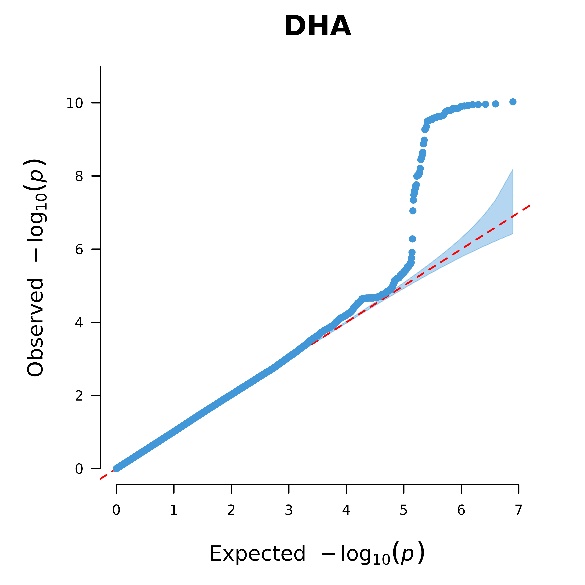

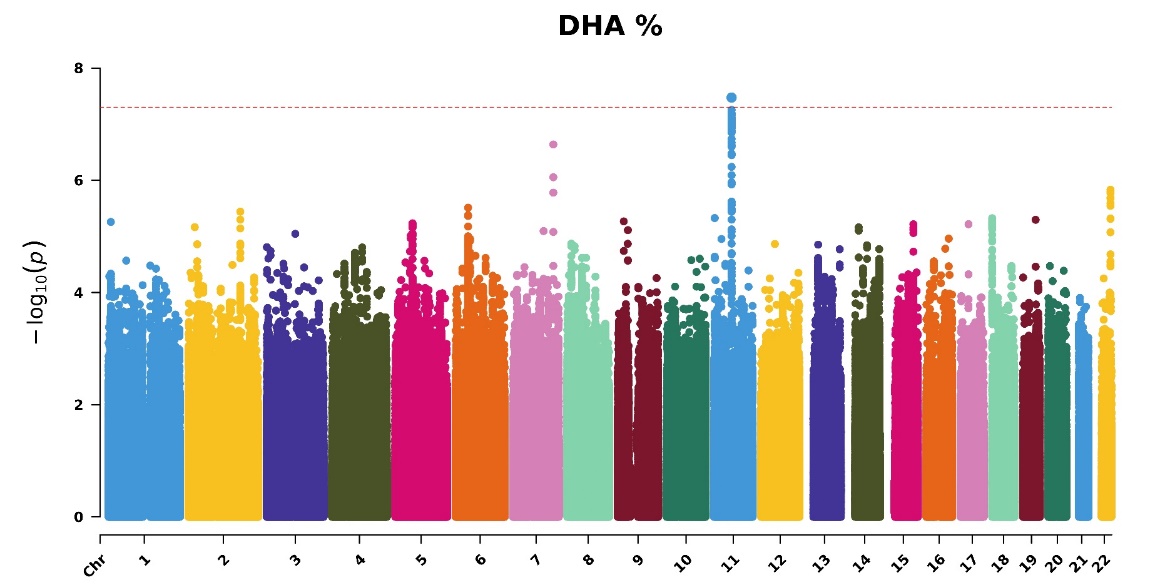

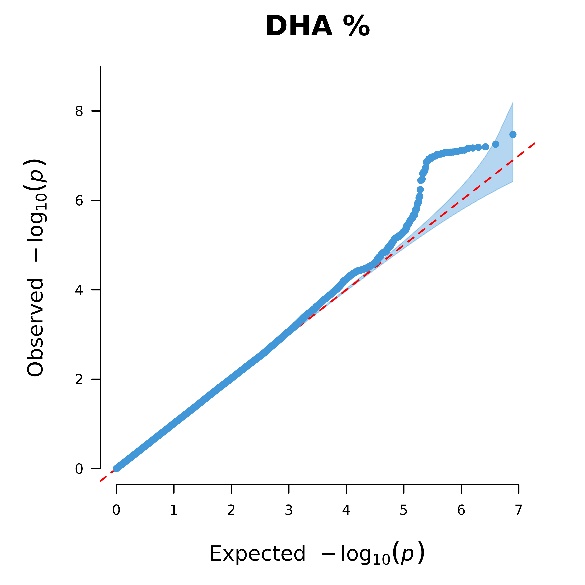

### Figure S4. Manhattan and QQ plots of *p*-values for gene-fish oil interactions in 14 PUFAs and MUFAs-related phenotypes for 114,352 participants in the Phase Two dataset

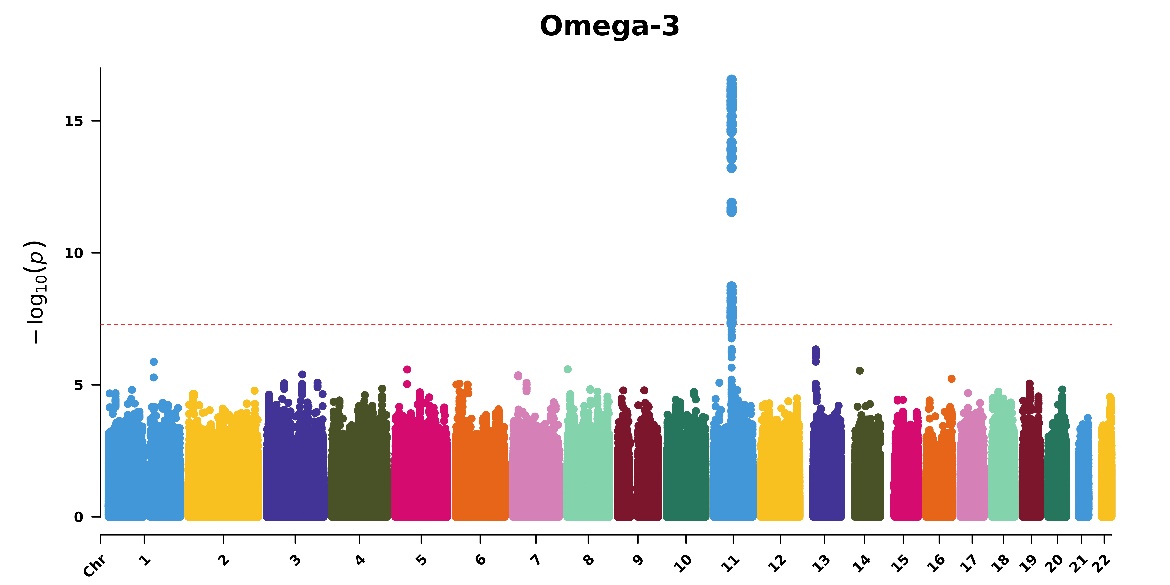

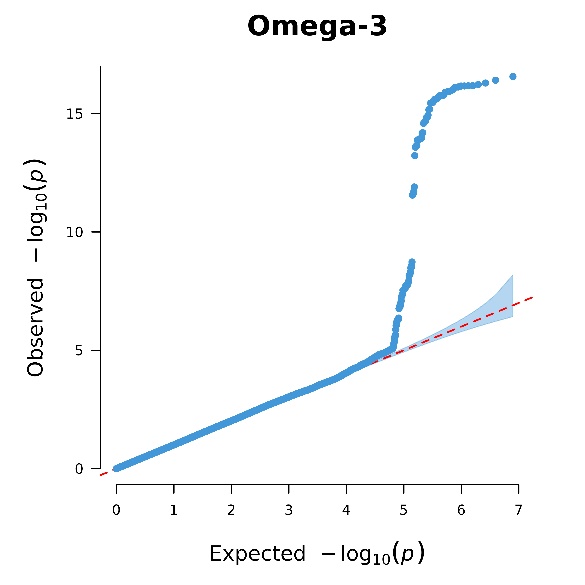

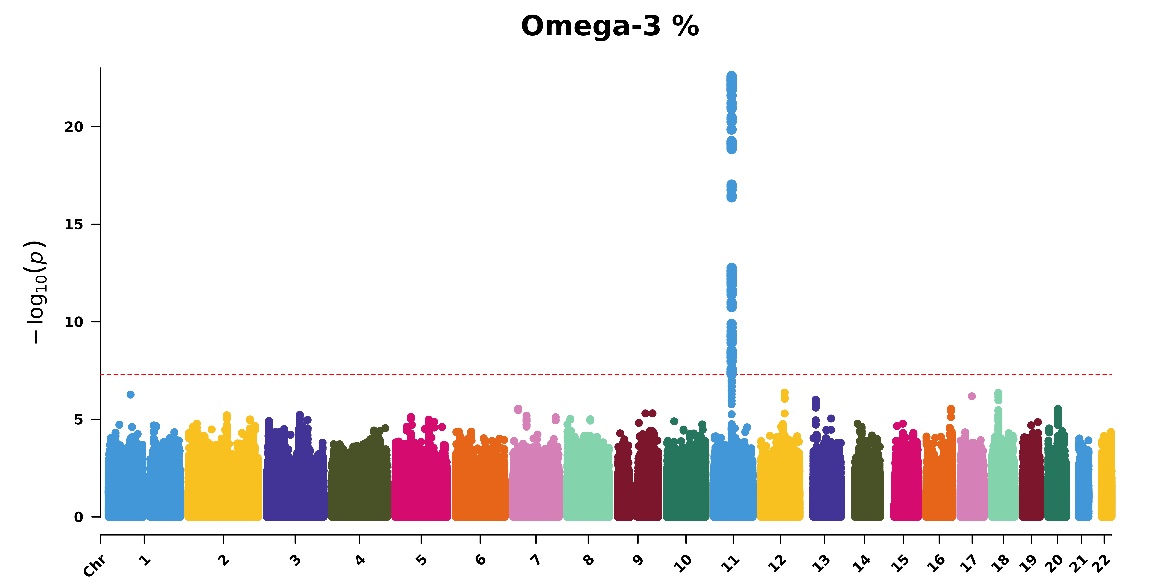

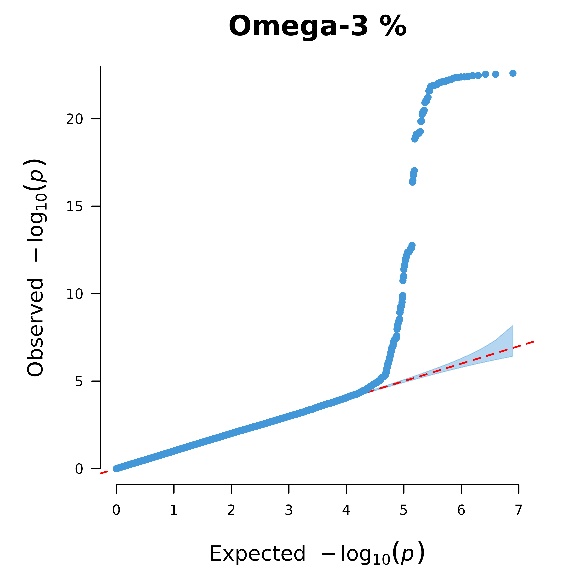

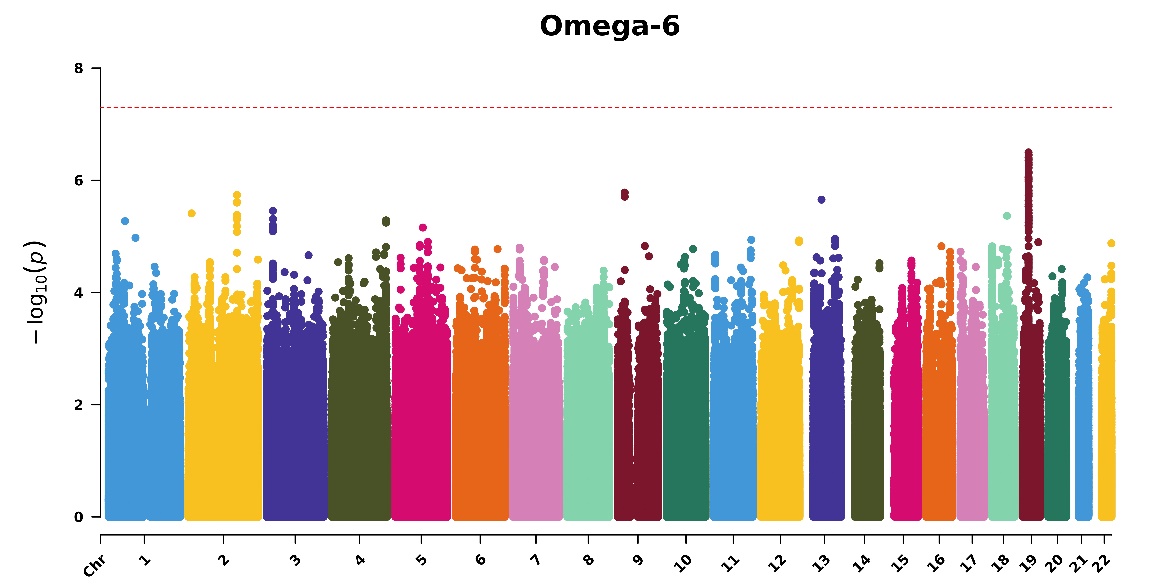

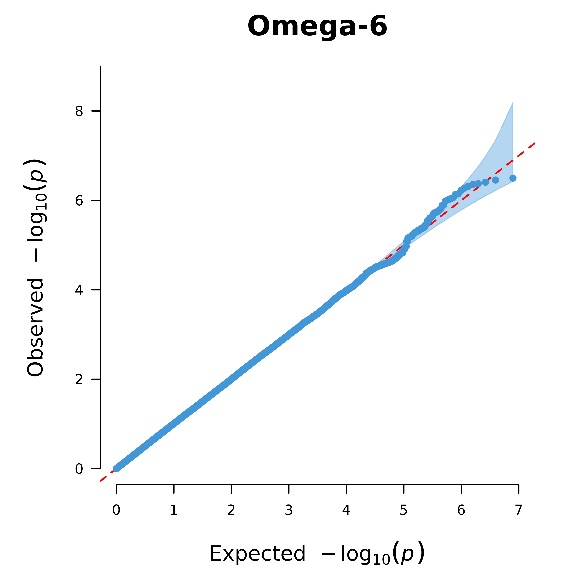

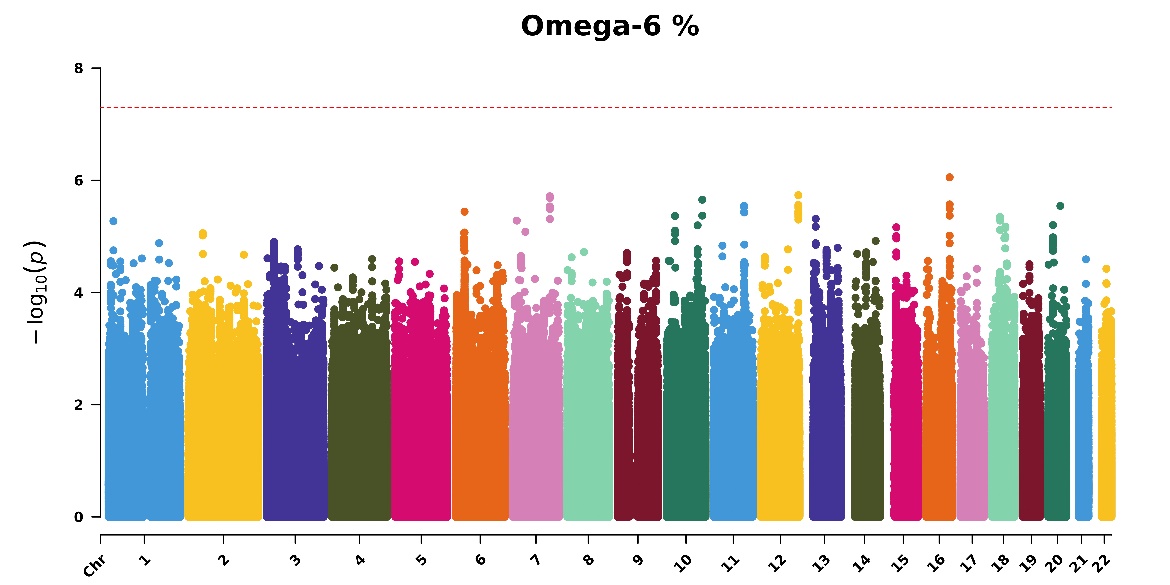

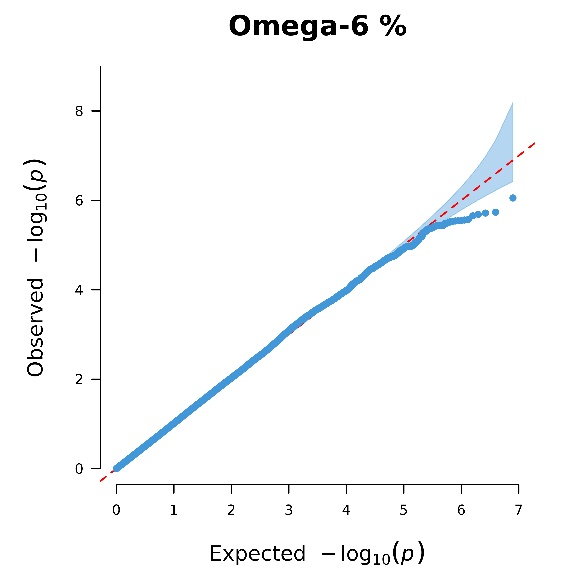

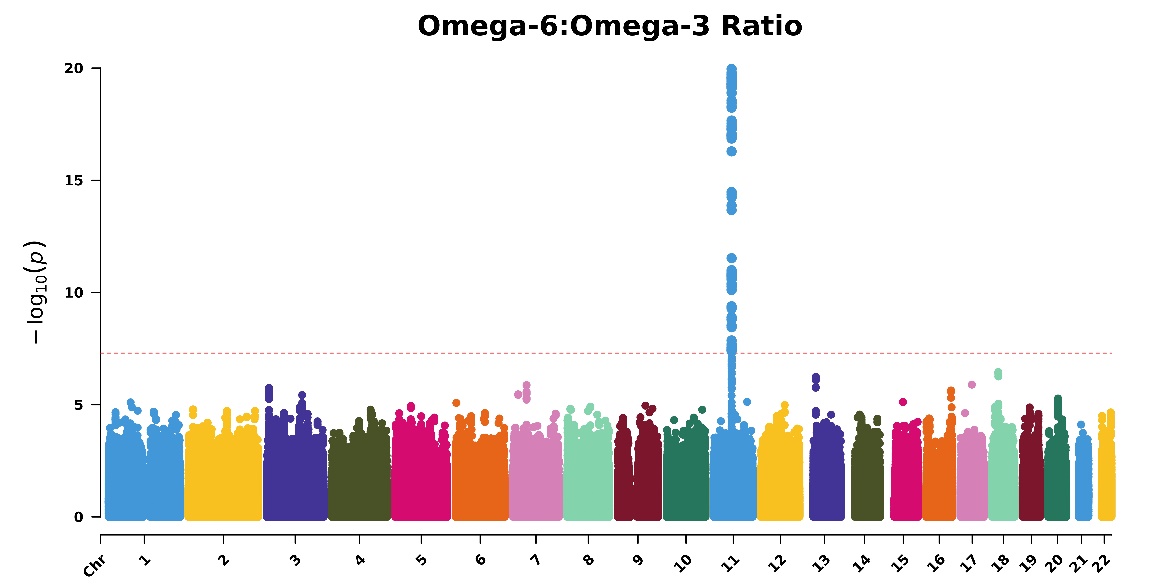

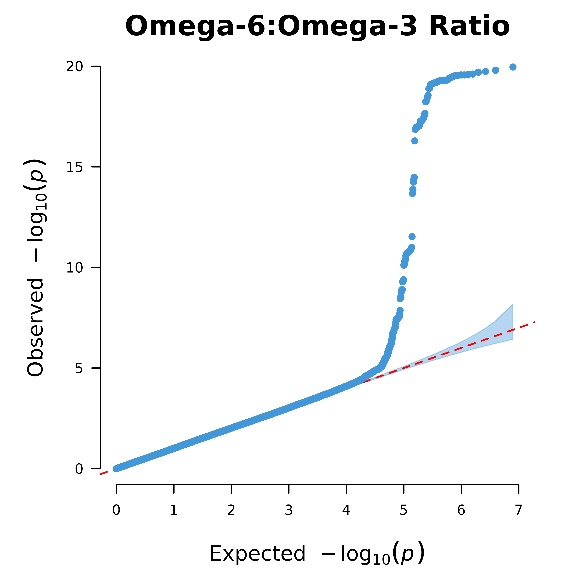

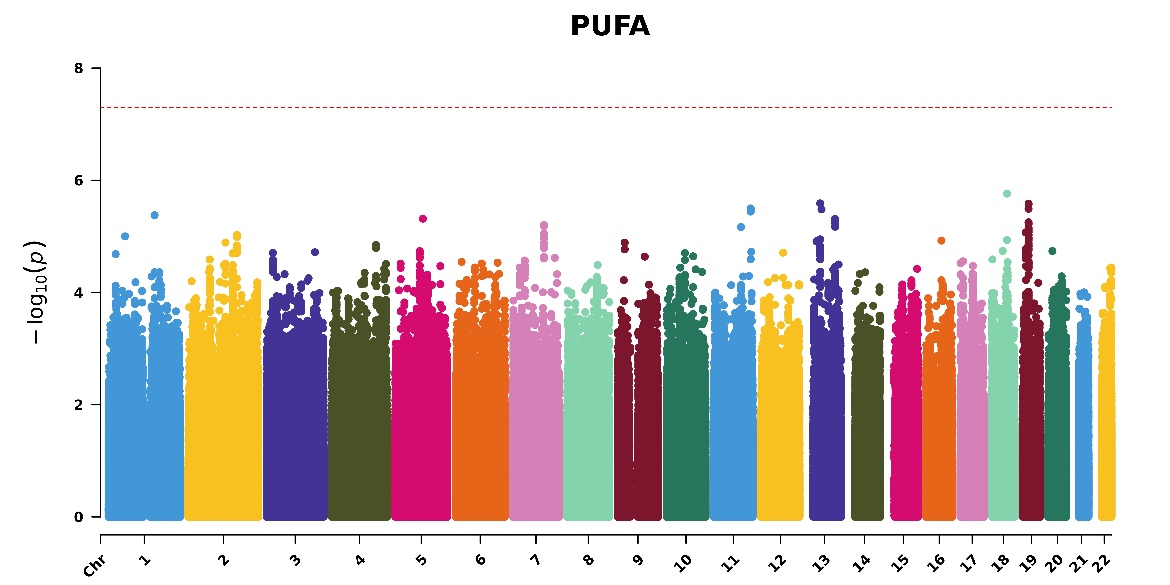

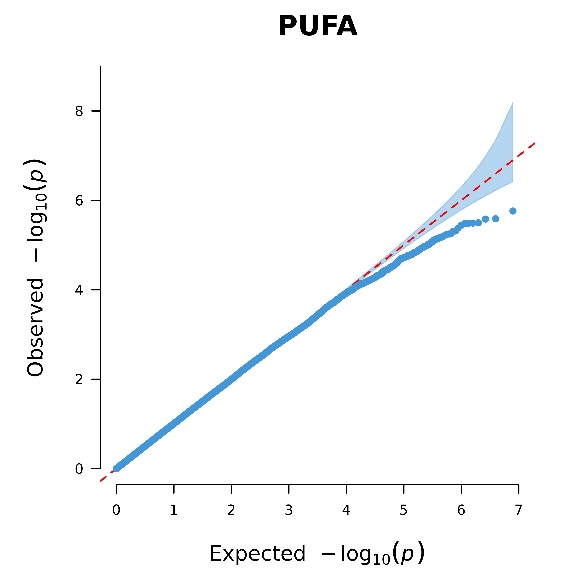

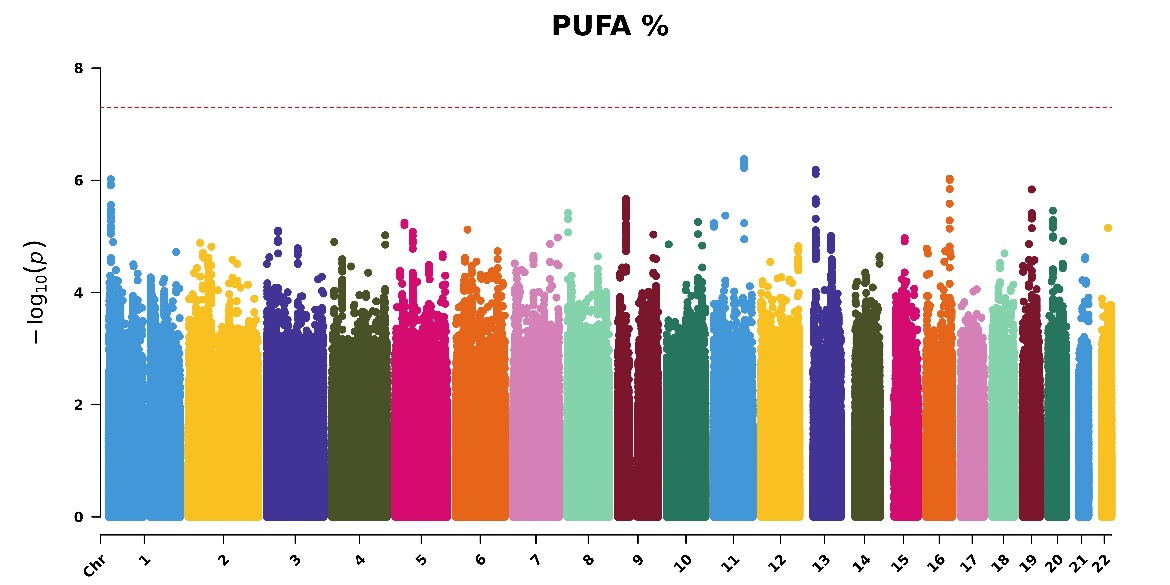

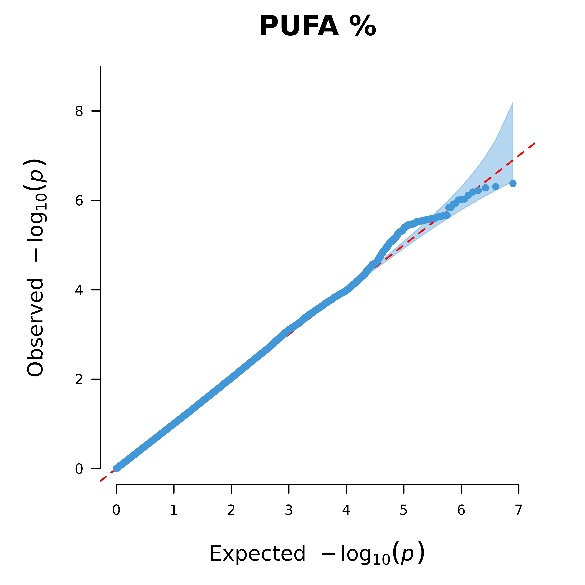

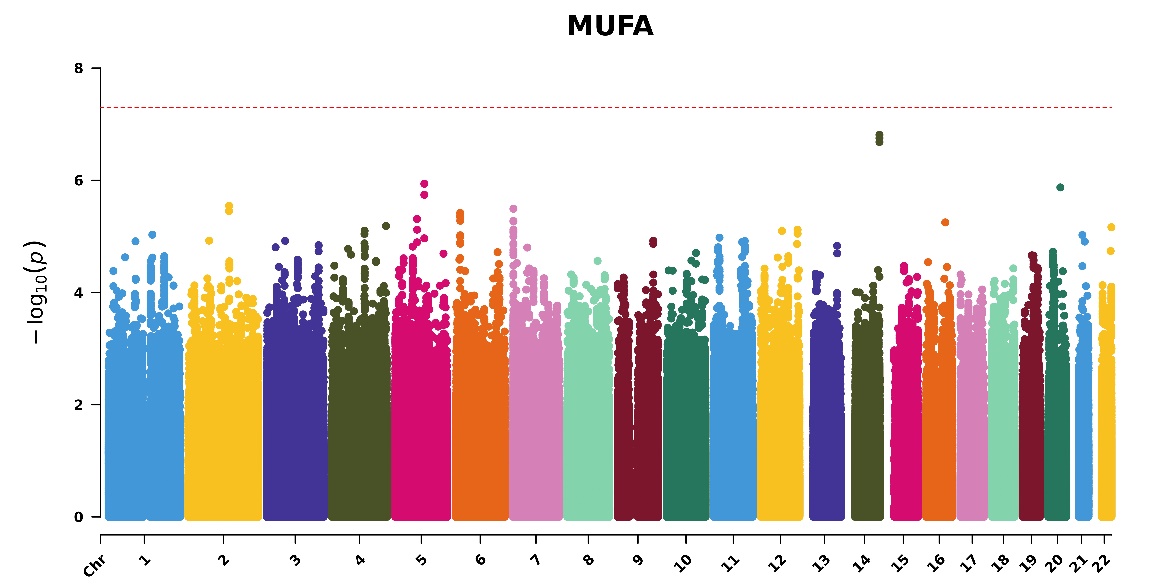

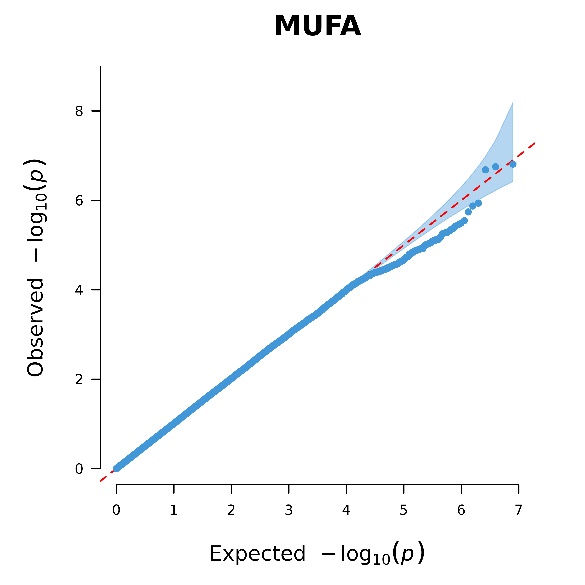

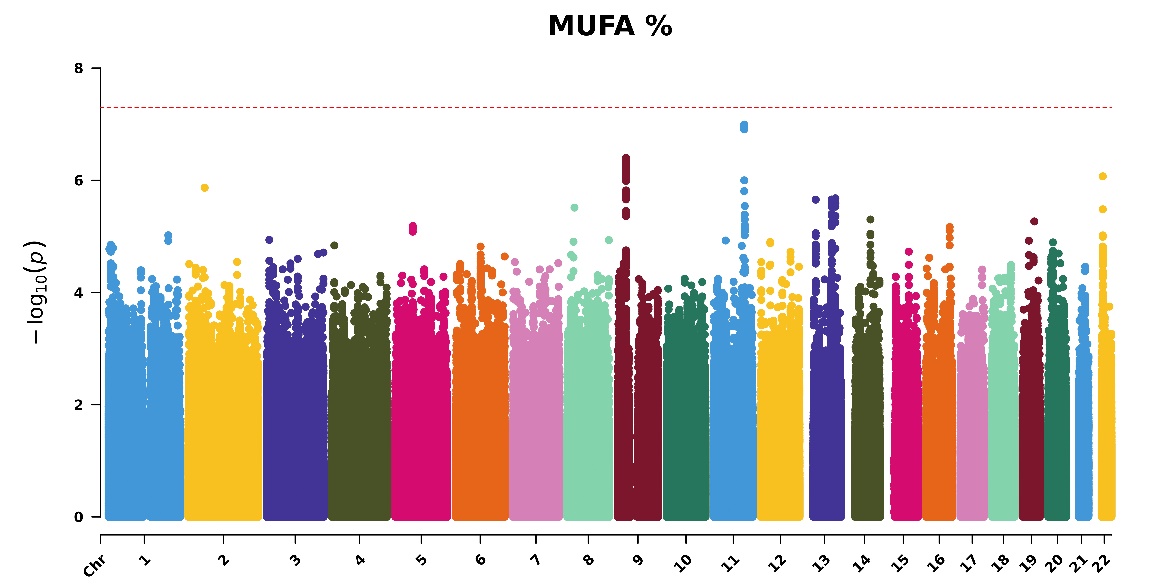

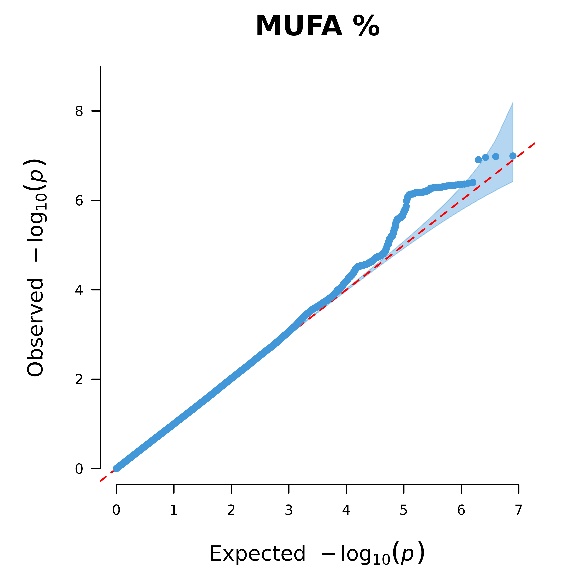

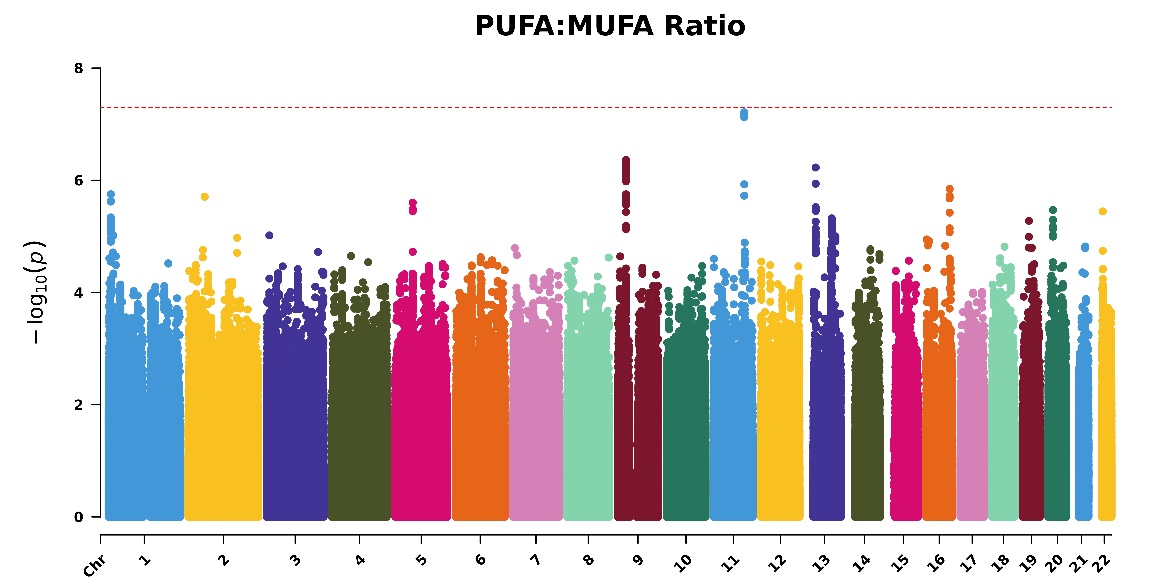

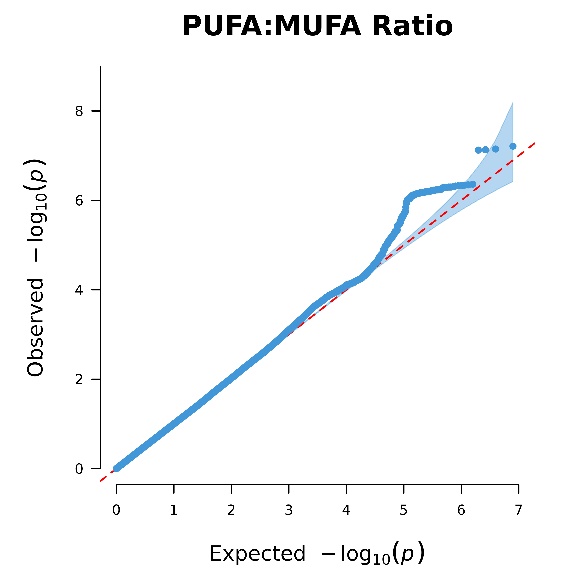

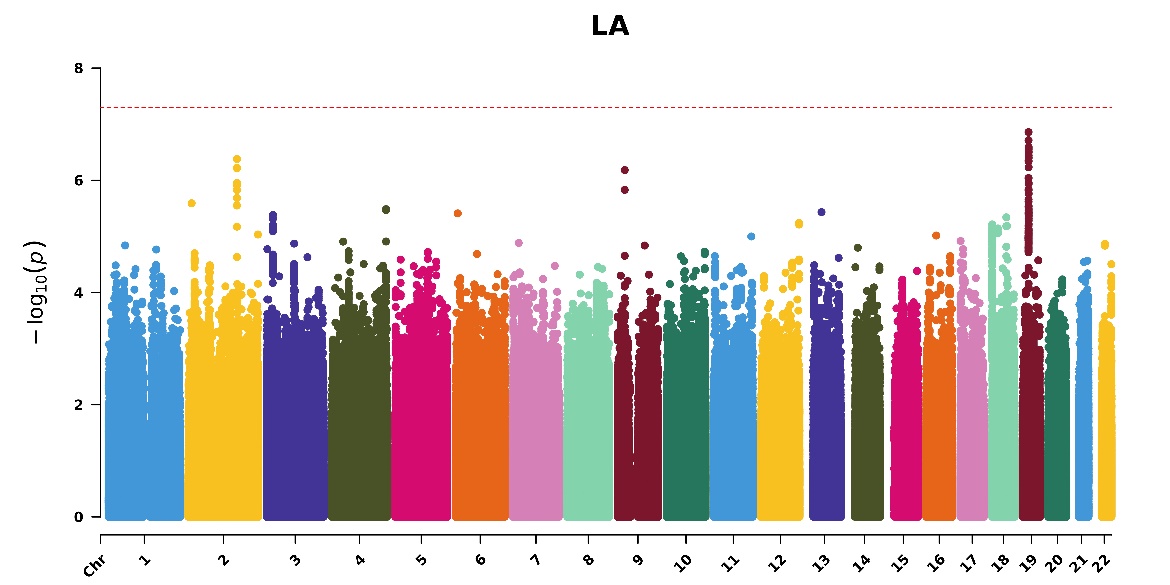

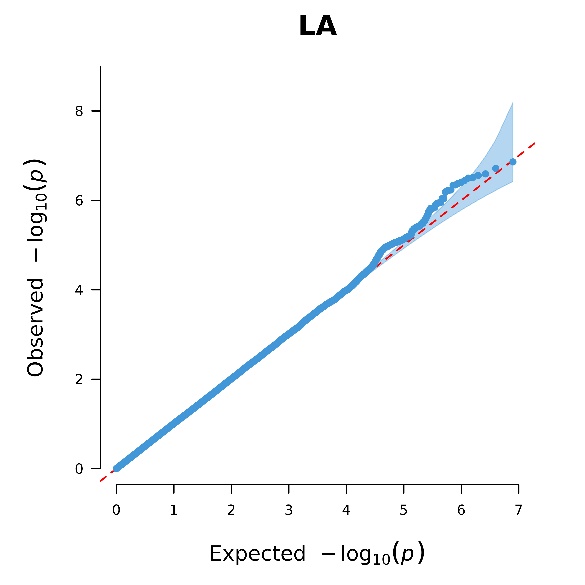

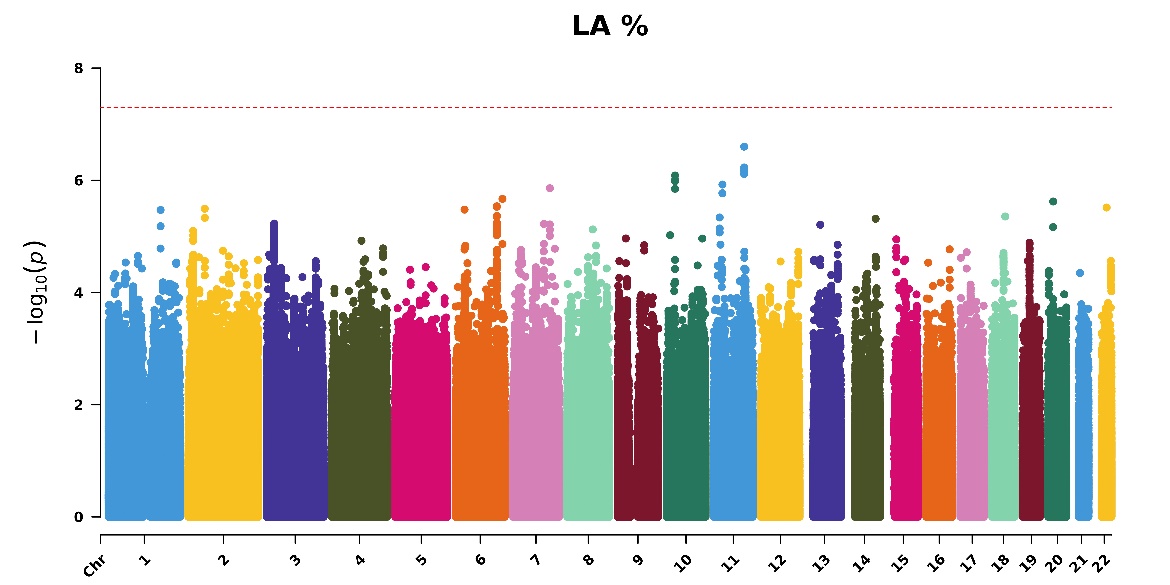

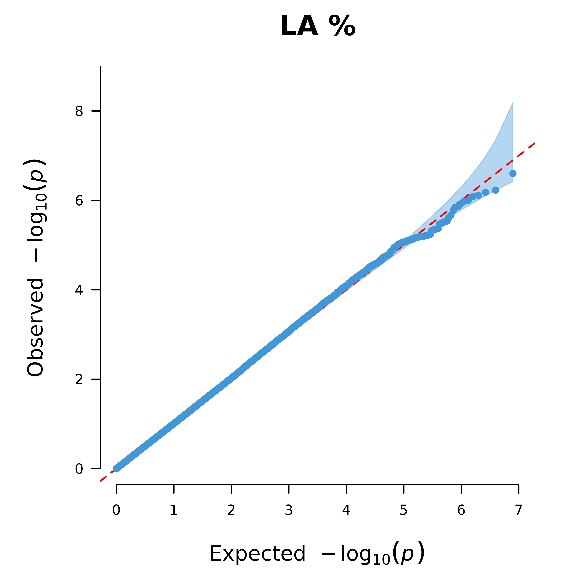

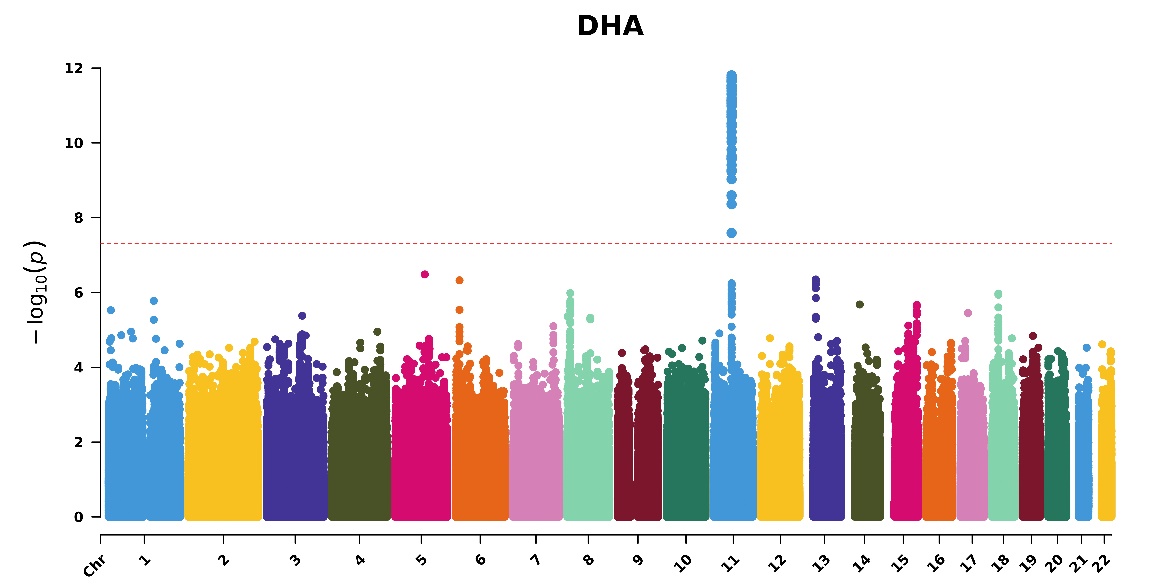

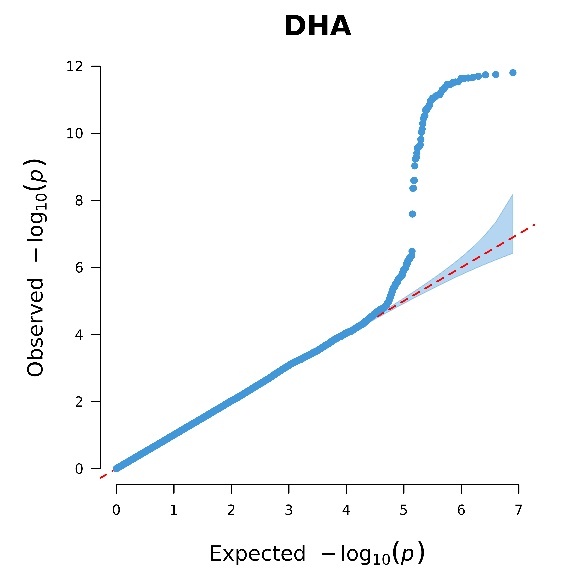

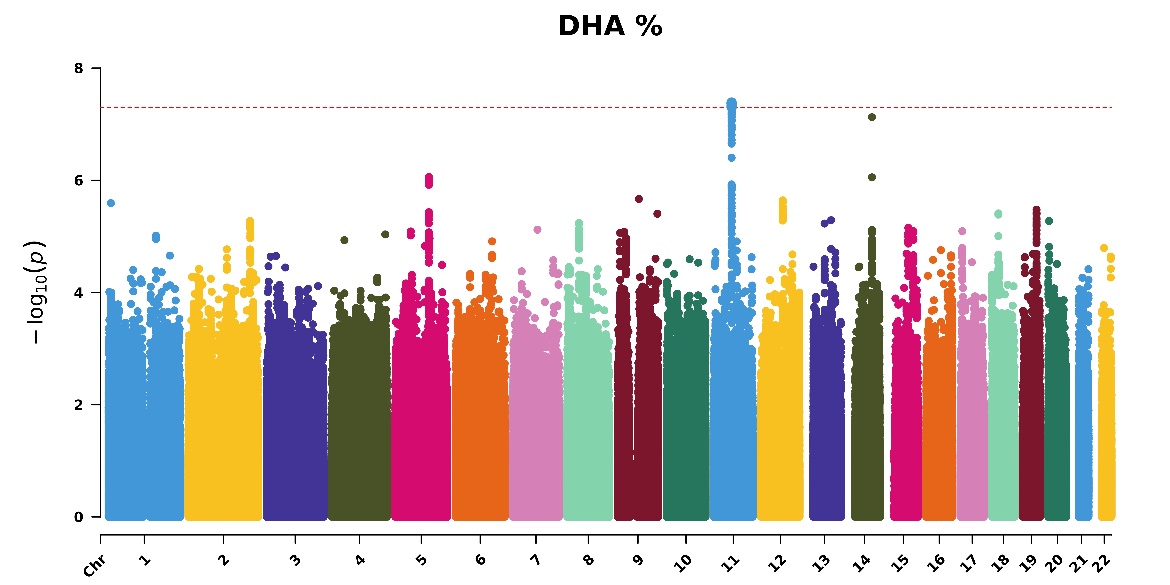

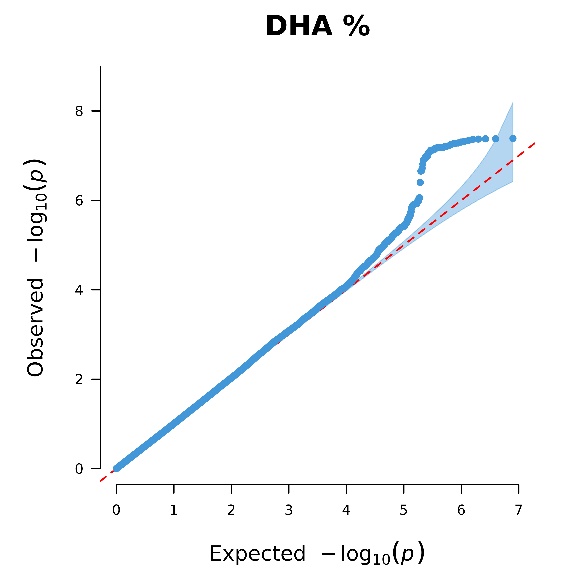

### Figure S5. Manhattan and QQ plots of *p*-values for gene-fish oil interactions in 14 PUFAs and MUFAs-related phenotypes for 200,060 participants in the combined dataset

### Table S1. Baseline characteristics of participants in Phase One, Phase Two, and combined releases of NMR metabolite data of individuals of European ancestry in UK Biobank

|  | Phase One  (*n* = 85,708) | Phase Two  (*n =* 114,352) | Combined phases  (*n =* 200,060) |
| --- | --- | --- | --- |
| Age, years (SD) | 57 (8.01) | 57 (8.00) | 57 (8.01) |
| Sex, female (%) | 45,907 (54) | 61,137 (54) | 107,044 (54) |
| Fish oil intake, *n* (%)  Yes  No |  | | |
|  | 27,201 (32) | 77,842 (32) | 63,711 (32) |
|  | 58,507 (68) | 36,510 (68) | 136,349 (68) |
| Absolute PUFA concentrations, mmoL/L (SD)  Omega-3  Omega-6  DHA  LA  MUFA  PUFA |  | | |
|  | 0.52 (0.22) | 0.54 (0.22) | 0.53 (0.22) |
|  | 4.44 (0.67) | 4.56 (0.69) | 4.51 (0.69) |
|  | 0.23 (0.082) | 0.24 (0.085) | 0.24 (0.083) |
|  | 3.41 (0.67) | 3.51 (0.69) | 3.46 (0.69) |
|  | 2.83 (0.81) | 2.98 (0.85) | 2.92 (0.84) |
|  | 4.97 (0.79) | 5.10 (0.82) | 5.04 (0.81) |
| Proportion in total fatty acids, % (SD) |  |  |  |
| Omega-3 | 4.39 (1.53) | 4.35 (1.53) | 4.37 (1.53) |
| Omega-6 | 37.97 (3.59) | 37.57 (3.58) | 37.74 (3.59) |
| DHA | 2.00 (0.66) | 1.98 (0.67) | 1.99 (0.67) |
| LA | 28.94 (3.40) | 28.71 (3.38) | 28.81 (3.39) |
| MUFA | 23.55 (2.67) | 23.97 (2.65) | 23.79 (2.67) |
| PUFA | 42.37 (3.73) | 41.92 (3.72) | 42.11 (3.73) |
| Ratios (SD) |  |  |  |
| Omega-6 to Omega-3 | 9.82 (4.28) | 9.81 (4.23) | 9.81 (4.25) |
| PUFA to MUFA | 1.84 (0.35) | 1.78 (0.33) | 1.81 (0.34) |
